# The National Institute on Aging Late Onset Alzheimer’s Disease Family Based Study: A Critical Component of the International Effort to Understand Alzheimer’s Disease

**DOI:** 10.1101/2021.04.12.21255332

**Authors:** Dolly Reyes-Dumeyer, Kelley Faber, Badri Vardarajan, Alison Goate, Alan Renton, Michael Chao, Brad Boeve, Carlos Cruchaga, Margaret Pericak-Vance, Jonathan L. Haines, Roger Rosenberg, Debby Tsuang, Robert A. Sweet, David A. Bennett, Robert S. Wilson, Tatiana Foroud, Richard Mayeux

## Abstract

**INTRODUCTION:** The National Institute on Aging Late-Onset Alzheimer’s Disease Family Based Study (NIA-LOAD FBS) was established to study the genetic etiology of Alzheimer’s disease (AD).

**METHODS:** Recruitment focused on families with two living affected siblings and a third first degree relative similar in age with or without dementia. Uniform assessments were completed, DNA was obtained as was neuropathology, when possible. *APOE* genotypes, genome-wide SNP arrays and sequencing was completed in the majority of families.

**RESULTS:** A wide range in the age-at-onset in many large families was related to *APOE* genotype, but not in all. Variants typically associated with early-onset AD and frontotemporal dementia were also found.

**DISCUSSION:** The NIA-LOAD FBS is the largest collection of familial AD worldwide, and data or samples have been included in 126 publications addressing the genetic etiology of AD. Genetic heterogeneity and variability in the age-at-onset provides opportunities to investigate the complexity of familial AD.

## 1. Introduction

In 2020, 5.8 million Americans were living with Alzheimer’s disease (AD). By 2050 this number is projected to grow to 13.9 million people, nearly 3.3% of the US population^1^. The etiology of AD remains unclear, but there is substantial evidence that AD is a highly heritable disorder. The National Institute on Aging Late-Onset Alzheimer’s Disease Family Based Study (NIA-LOAD FBS) is the largest collection of multiplex AD families recruited and longitudinally assessed worldwide. Since its inception in 2003, the central goal of this cohort was to broadly support genetic research by making clinical and genomic data and biological samples broadly and rapidly available to the AD research community to support the development of therapies to treat or prevent this disease. The use of multiplex families to understand genetic susceptibility to AD began with early onset forms. These studies led to the discovery of rare and highly-penetrant mutations in autosomal dominant AD: *APP*^*2*^, *PSEN1*^3,4^and *PSEN2*^*5,6*^. Nearly 60% of individuals with early onset AD have other affected family members,^7^ and of these, approximately 13% inherit these mutations in an autosomal dominant manner^8^. While variants in these genes have explained only a small proportion of the occurrence of familial, early-onset AD, they formed the basis for the development of novel therapies now being investigated in patients with AD regardless of age at onset of disease^9^.

Genetic research on the more common late onset form of AD was transformed by the work of Pericak-Vance and her colleagues^10^ who used a novel linkage strategy in multiply affected families with late-onset AD. They were the first to identify a locus on chromosome 19^10^, which subsequently lead to the discovery of the association between the *APOE* ε4 allele and the increased risk of developing AD and the association of the *APOE ε2* allele and reduced risk ^11,12^. The association was later confirmed in almost all ethnic groups, but with a slightly weaker effect among those with African ancestry^13^. Overall, the population attributable risk of the *APOE-ε4* allele has been estimated at 20%^14^. Nonetheless, the accumulating evidence indicates that late-onset AD is multifactorial with high heritability, 70% or higher, implying that other genes or common risk factors may also contribute to its etiology.

Genetic investigations typically require time and effort to ascertain samples for genetic studies. While it was presumed that recruitment of unrelated patients and controls would be a faster and more efficient way to find risk alleles than families, there are many scientific and practical advantages for recruiting families for genetic research. The observed incidence rates for AD has been estimated to be three to five times higher in multiplex families^15^. Multiplex families may be enriched in disease causing genetic variants, allowing penetrance to be studied. This familial enrichment is particularly valuable for rare variants (MAF<0.01), where population-level studies have little power. Family members are generally eager to participate in such research and often have shared environments and parents of origin. Moreover, they also provide the opportunity to assess interaction between alleles at the same locus and to detect epistasis due to the high penetrance.

The NIA-LOAD FBS has contributed to nearly every genetic study of AD and with the advent of novel strategies to understand how genetic variation leads to this disease, it will continue to do so. Here we describe the cohort in detail, show the range of age-at-onset across and within families, the effects of *APOE* on the risk of AD across ethnic groups, the genetic heterogeneity and highlight the broad availability of clinical and genetic data and biological samples.

## 2. Methods

### 2.1 Participant recruitment

Families assumed to be multiply affected by AD by history or autopsy were recruited from sites across the United States (supplemental figure 1). The criteria for study entry included two living affected siblings, 60 years of age or older, and a third living relative, similar in age, with or without dementia. We also included families with deceased affected siblings, as long as frozen brain tissue was available for affected members. Families in which participants were symptomatic, but did not meet criteria for AD, were still included with the commitment for follow-up. Healthy controls without a family history of AD in a 1^st^ degree relative, who were 55 years of age or older were also recruited. Participants demonstrated the full range of cognitive function including normal cognition and dementia (table 1). All participants considered to have clinical AD were required to meet established research criteria the disease^16^.

Families were enrolled and consented at individual study sites and completed the following protocols either by in person or telephone interview and assessments. The study protocol included: a family pedigree, standardized cognitive and psychiatric assessments, as well as a review of AD and cardiovascular risk factors (table 1). Regular follow-up visits were used to update the family history and obtain additional clinical information on new family members that had not previously participated. All study data were stored at the Columbia University, the Coordinating Center.

### 2.2 Cognitive assessment

Standardized cognitive assessments began in 2006, and 75% of the cohort has been assessed^17,18^. The 15-minute cognitive battery consisted of seven brief tests that were administered in person or by telephone and assessed: Working memory - Digit Span Forward and Backward and Digit Ordering; Episodic memory - Immediate and Delayed Recall of story “A” from the Wechsler Memory Scale and Semantic memory - asked persons to name members of two semantic categories (Animals, Vegetables) in separate 1-minute trials. From these measures, a factor analysis of the seven tests summary measures of working memory, declarative memory, episodic memory, semantic memory, and global cognition were developed. The interviews using different methods, in-person or by telephone, had little effect on performance and provided operational flexibility and a means of reducing costs and missing data^17^.

### 2.3 Psychiatric Interview

The assessment of psychiatric manifestations was added in 2010 and was designed to be completed in an interview with a knowledgeable informant and could be conducted after the participant’s death. This approach captured psychotic symptoms that occurred in the prior month, identifying the month during which each symptom was most persistent and recording the frequency of symptoms^19^. The Neuropsychiatric Inventory (NPI-Q)^19,20^, a widely used, validated informant-based interview that is used to evaluate neuropsychiatric symptoms in patients with dementia was selected. The NPI-Q has good interrater reliability and concurrent validity compared with more structured interviews^21,22^.

### 2.4 Antecedent risk factors

We collected putative risk and protective factors associated with AD, and at follow-up we reviewed the previous risk factor profiles to update any changes. The primary focus was on modifying factors such as hypertension, heart diseases (myocardial infarction, congestive heart failure, or other type of heart disease) and diabetes^23^. A stroke assessment captured the number of stroke events. The structured interview concerning modifying antecedent (risk or protective) factors was also used in follow-up by telephone. For participants unable to be interviewed, the person who best knew the patient provided the information. Other factors to be ascertained included history of head injury, smoking, alcohol use, physical activities and leisure activities. Current medications were queried. In addition, for each medical condition or risk factor endorsed, the age at onset, duration, treatment history and whether there had been any change in the condition since the prior assessment was ascertained.

### 2.5 Biological Resources Collection

Initially, blood, saliva or frozen brain tissue collected at baseline and follow-up visits were sent to the National Centralized Repository for Alzheimer’s Disease and Related Dementias (NCRAD) which extracted and stored DNA. As shown in supplemental table 1, we collected DNA from 7,925 participants. In 2018, we expanded blood collection to also obtain peripheral blood mononuclear cells (PBMCs) at follow-up visits.

### 2.6 Genotyping and sequencing

Supplemental table 1 summarizes available genetic data. Genome wide SNP array (GWAS) data was obtained in 1,340 families (76.3%) in the cohort, which included 7,012 (72.4%) of their family members. Whole exome (WES) or whole genome sequencing (WGS) was conducted in 1,340 (76.3%) of the families. *APOE* genotyping was performed in 1,260 families (71.8%). The results of all genetic studies have been stored at the National Institute on Aging Genetics of Alzheimer’s Disease Storage Site (NIAGADS).

### 2.7 Neuropathological confirmation of diagnosis

At the inception of the study, NIA-LOAD FBS began preplanning brain donation with all family members to maximize the likelihood that neuropathology would be performed after death. This resulted in a robust brain donation program in which autopsy reports or brain tissue was successfully obtained in 836 individuals (table 2). In the event an autopsy was performed, and no tissue was available, the autopsy report was requested. Brain autopsy was performed at Columbia University or at one of the study sites, with diagnoses based on the 2012 NIA-Alzheimer’s Association Working Group guidelines^24^. Brain tissue, frozen and fixed, were sent to NCRAD to share with the research community. In 2019 we expanded the use of brain tissue to serve not only as a means to confirm diagnosis, but also to support functional studies including transcriptomics and DNA methylation studies.

## 3. Results

### 3.1 Demographics

Through a national effort, we recruited 1,756 families and acquired data from 9,682 family members. The families were diverse: 181(10.3%) African American, 425 (24.2%) Caribbean Hispanic, 138 (7.8%) self-reporting as “other or mixed” and 1,012 white, non-Hispanic (supplemental table 2). Among the affected and unaffected family members, 63% were women, a percentage that differed significantly by ethnic group (72% among African Americans and 66% among Hispanics, 62% among white non-Hispanics and 57% the other group, p=0.009). Among the 1,756 families, 451 (25.7%) had three or more affected family members. Another 31.5% had two affected individuals while 598 (34%) families consisted of only a single affected individual or a single individual who, after clinical evaluation, did not meet criteria for dementia or AD but had cognitive impairment. These families were still included for longitudinal observation. The proportions of affected to unaffected individuals was similar across the ethnic groups (supplemental table 2).

### 3.2 Data collection

In the family cohort, 8,610 (77.7%) individuals had detailed clinical information including multiple cognitive assessments. The other 22.3% were either too impaired for cognitive testing or were recruited before the clinical assessments were standardized in 2007. Tables 1 and 3 summarize the data collected and the resulting clinical diagnoses. Due to the national recruitment of families, we initiated telephone interviews several years ago to make it easier for family members to continue to participate. We conducted in-person interviews in 7,412 (68.5%) and follow-up interviews in 3,349 (30.9%). Because we had alternatives to in-person assessment in place prior to the pandemic, we were able to continue to follow family members; however, our ability to recruit new families was limited. Due to the length of this study, nearly 30 years for some families, 15.1% of the cohort has been lost to follow-up without leaving a forwarding address or telephone number, and 2,878 (26.6%) died.

### 3.3 Age-at-onset

The average age of onset in affected family members differed slightly, but significantly, across all ethnic groups ranging from 72 years in Caribbean Hispanics to 74 years in African Americans and white non-Hispanics (p<0.0001). The average age at last visit among unaffected family members differed significantly by ethnic group (66.2 yrs, African American, 63.9 yrs, Hispanic vs. 68.7 yrs, white non-Hispanic; p<0.0001).

We observed a very wide range of age-at-onset among the families with three or more affected members (figure 1). To explore the variation in onset age further we investigated families in which some individuals had their age-at-onset at least 10 or more years later than the mean for the family (figure 1b). Among these 132 families, the average age at onset was 71.79 years. Individuals with later ages-at-onset were significantly less likely to carry an *APOE-ε4* allele (44.9% vs. 60.5%) and more likely to carry an APOE-ε3 allele (47.1% vs. 33.3%).

**Figure 1a.**
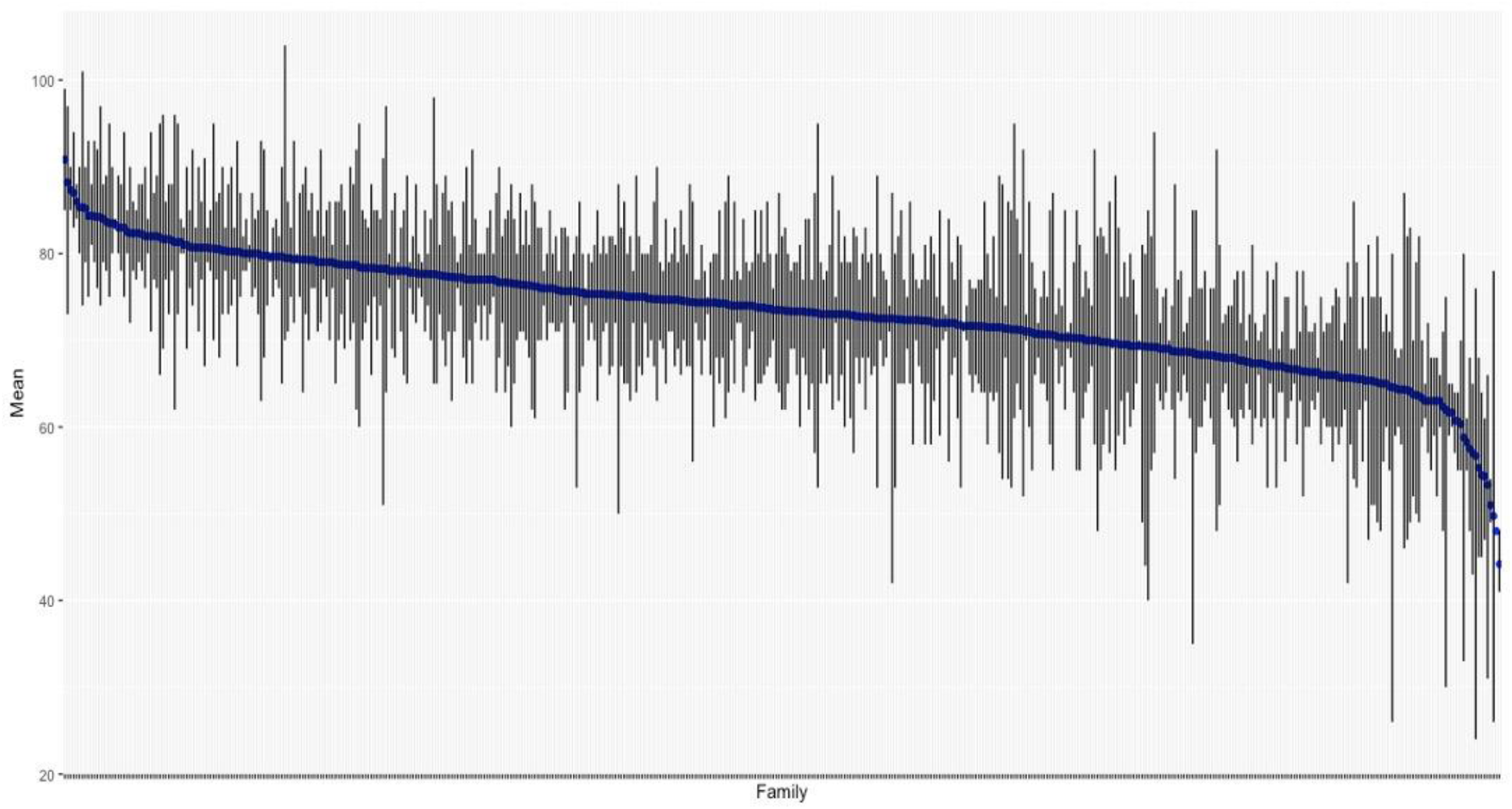
Distribution of age-at-onset among 451 families with three or more members affected by AD. The blue horizontal line represents the mean age at onset for each family, while the gray vertical lines represent the range of onset ages within each family.

**Figure 1b.**
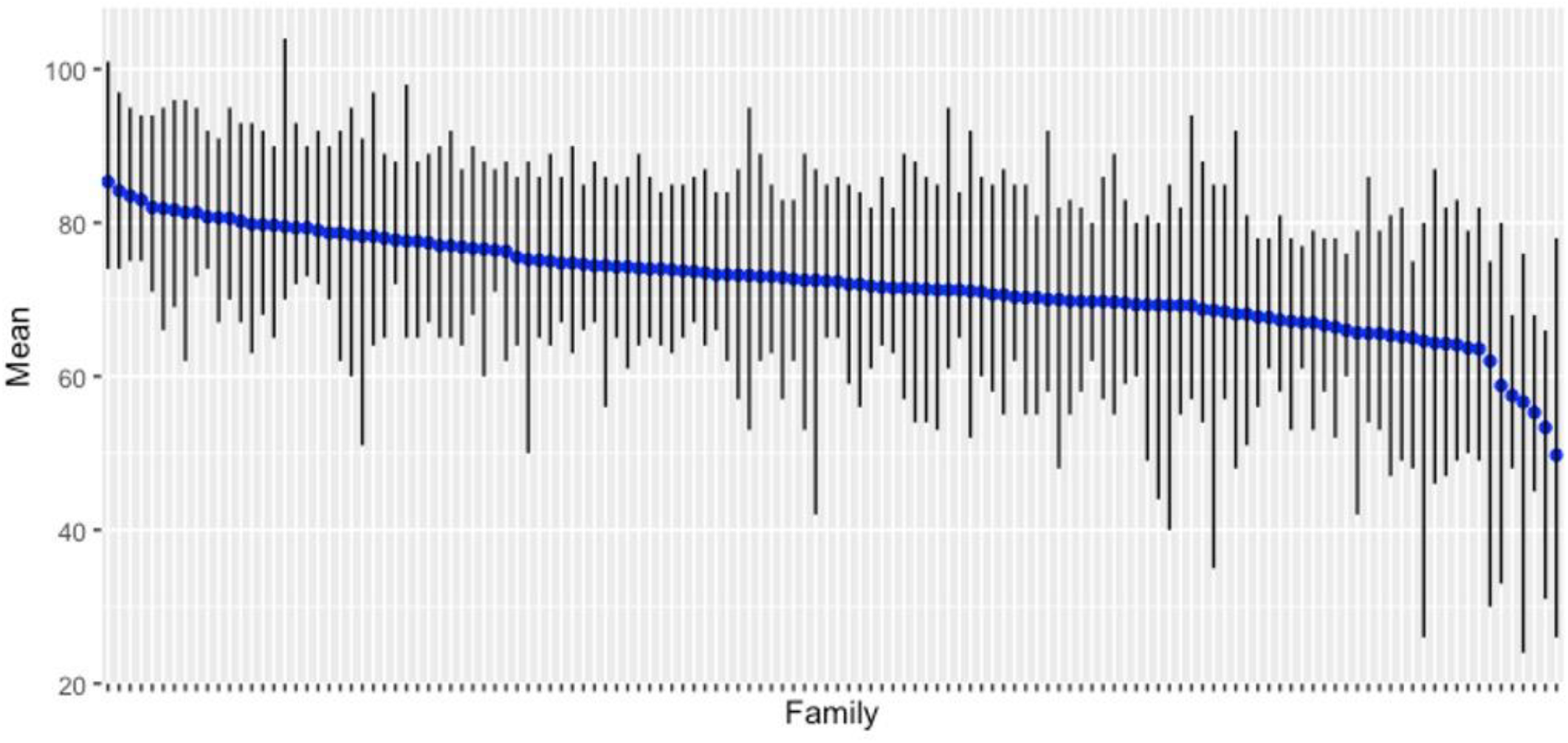
Distribution of age-at-onset among 132 families with three or more members affected by AD and with a delay in onset by 10 years or more from the mean age-at-onset in the family. The blue horizontal line represents the mean age at onset for each family, while the gray vertical lines represent the range of onset ages within each family.

### 3.4 Genotyping

*APOE* genotyping revealed significant effects on the age-at-onset across all families, with similar effects in women and men (supplemental figures 2a and 2b). The *APOE-ε4* allele had the strongest effects in white, non-Hispanic families when the genotype was homozygous or heterozygous. However, among Caribbean Hispanics and African Americans the genotypic effects were limited to *APOE-ε4* homozygotes (supplemental figures 3a-d). These studies indicate that although *APOE* is a strong risk factor for sporadic AD, and also plays a role in familial AD.

**Figure 2.**
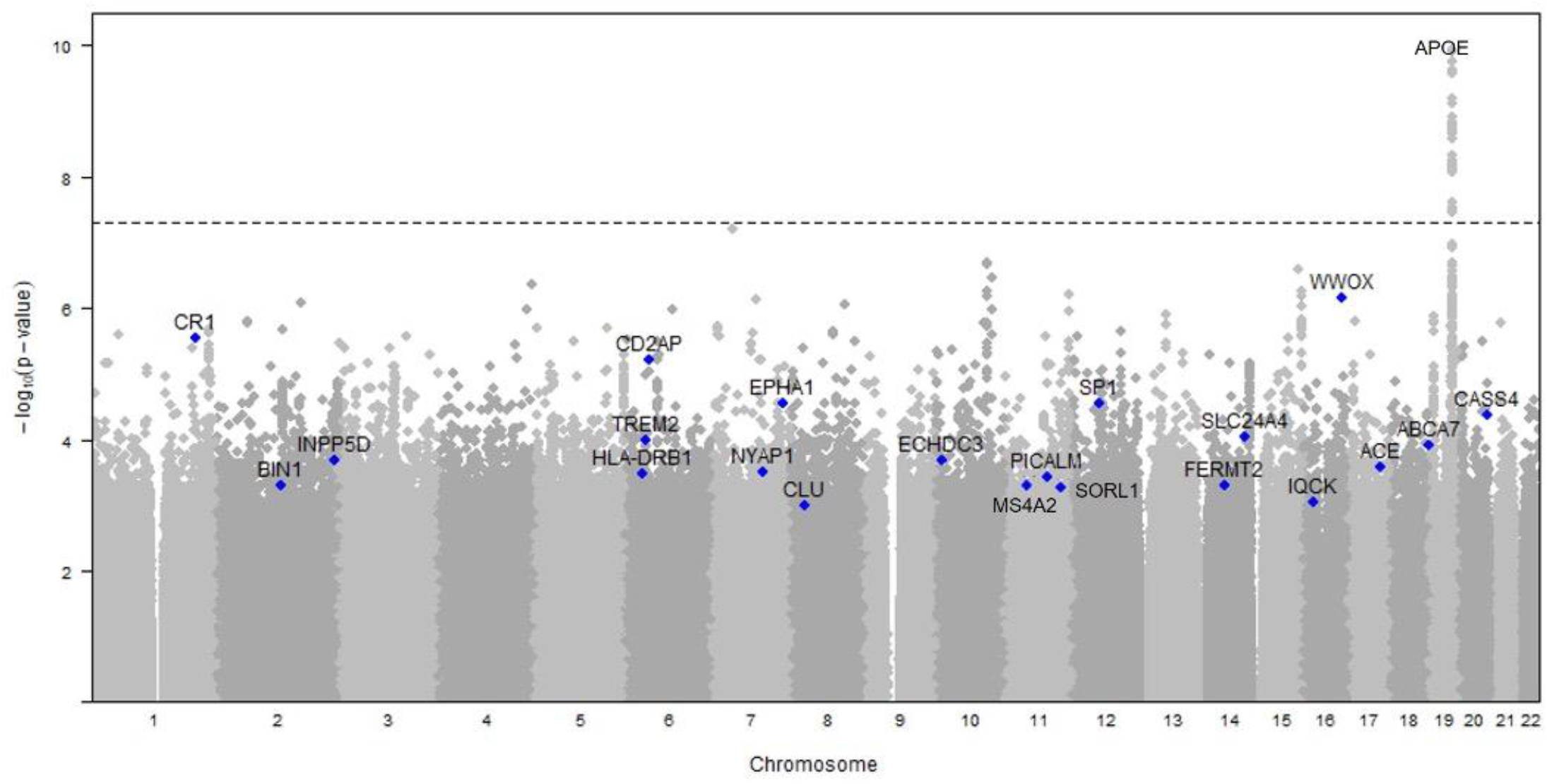
Results of genome-wide array in the NIA-LOAD FBS. The APOE locus reached genome-wide significance in associated with AD. However, a number of other loci, labeled on the Manhattan plot below were nominally significant. Many of these loci supported several large meta-analyses that included other cohorts^32-34^.

#### 3.4.1 Pathogenic variants

We found variants in *APP, PSEN1 and PSEN2* associated with familial early onset forms of AD and *MAPT, GRN* and *C9orf72* with early onset forms of frontotemporal dementia (FTD). Sequencing these families have demonstrated that pathogenic variants in these genes can be also found in late-onset families ^25-29^. The two more common pathogenic variants found in these families are the PSEN1 p.A79V (0.46% of the families) and the C9ORF72 expansion (0.48% of the families). Among the probands, 1.82% carried pathogenic variants in *APP, PSEN1, and PSEN2*, and 1.94% had pathogenic variants in *MAPT, GRN*, or *C9orf72*. These findings also imply that modifying factors may exist, either genetic or environmental, that delay the onset of disease in these late-onset families. Therefore, additional studies in the NIA-LOAD FBS are warranted to identify these novel modifier or protective variants.

Finding pathogenic variants usually associated with FTD among families with an AD phenotype could either indicate misdiagnosis or pleiotropy. Although autopsies were not available for all the individuals with FTD pathogenic variants, the neuropathology in some individuals, was compatible with a diagnosis of AD with mixed pathologies^30^. *C9orf72* expansion has also been associated with several clinical presentations (FTD and ALS). The NIA-LOAD FBS extends these data to other genes and diseases, supporting the previous finding of pathogenic variants in FTD genes among individuals with AD pathology^31^.

### 3.5 Genomewide SNP Array

Despite the relatively modest size of the NIA-LOAD FBS data set, nominal significance in several loci, such as *CR1, CD2AP* and *EPHA1* contributed to several large AD meta-analyses (figure 2)^32-34^. The NIA-LOAD FBS dataset is also instrumental in the identification of additional risk variants. Recent students using polygenic risk scores constructed based on summary statistics for disease risk for sporadic AD demonstrated that these families are enriched in genetic risk factors^23,35^.

### 3.6 Neuropathological confirmation of diagnosis

In 492 (82%) participants, neuropathology confirmed the diagnosis of AD. Other diagnoses accounted for the remaining 163 with and without dementia (table 2). Brain tissue is available by request to NCRAD.

## 4. Discussion

The NIA-LOAD FBS began in 2003, with the collection and longitudinal follow up of large families multiply affected with AD. 1,885 DNA samples from 422 families were collected in the first three years from the NIA-funded AD Centers (ADC) throughout the United States. Subsequently, a goal was set to increase the collection to 1,000 families or more, to recruit all appropriate members in existing families, and initiate and complete follow-up. Blood samples from the participants were sent to the National Centralized Repository for Alzheimer’s Disease and Related Dementias (NCRAD) at Indiana University, for DNA extraction and storage allowing samples to be broadly available to the research community. Initially, clinical data regarding the diagnoses was limited to the criteria used by each Alzheimer’s Disease Center, but in 2007 a more standardized approach was used to collect clinical information in a uniform fashion, to store it centrally at the Columbia University Coordinating Center and to share it along with genetic data with the AD research community through NIAGADS. Nineteen dataset accession numbers contain genetic data from the NIA-LOAD FBS (table 4).

A genome wide association study (GWAS) was performed in the first 500 families, and data were deposited into the Database of Genotypes and Phenotypes (dbGaP) for broad sharing with the genetics community^36^. In 2012, the Alzheimer’s Disease Sequencing Project (ADSP) included the sequencing of 583 participants from 111 multiply affected families from the NIA-LOAD FBS^29,37^. The ADSP Discovery Extension Study (2015-2018) included whole genome sequencing (WGS) on individuals from multiply affected families provided by the NIA-LOAD FBS. The ADSP Follow-Up Phase (2018-2023) heavily engaged resources from the NIA-LOAD FBS and depended on this project to conduct continued longitudinal follow up of families and the further collection of 500 additional families from under-represented populations. The characterization of additional relatives, the ascertainment of antecedent risk factors and the recruitment for autopsy benefits the research community by expanding the scientific value of the NIA-LOAD FBS. Thus, over the past two decades, the NIA-LOAD FBS has acted as the key source of biological materials, genetic and longitudinal clinical data on large multiply affected AD families from ethnically diverse populations.

The range of age-at-onset varied within and across families is shown in figure 1a and 1b. We hypothesize that individuals whose age-at-onset of AD is more than 10 years from the mean within their family may have benefited from an as yet unidentified genetic or epidemiologic protective factor. In fact, these individuals were significantly less like to harbor an *APOE-ε4* allele and more likely to have an *APOE-ε3* or *-ε2* allele. Similarly, those individuals whose age-at-onset was 10 years earlier than the mean within their family may harbor additional genetic or environmental risk factors. Phenotypic variation among members of a family with dementia, including differences in the age of onset and the effects of *APOE* or the presence of rapid eye movement (REM) sleep behavior disorder, psychosis and parkinsonism may be due to interactions with other genes or epigenetic effects. Brain transcriptomics and DNA methylation may help to clarify the origin of these differences. Moreover, because vascular risk factors clearly contribute to the pathogenesis of AD^23^, the effects of cerebrovascular pathology remain to be defined.

As expected, the effects of *APOE* on the age-at-onset were statistically significant especially among white, non-Hispanics. In the homozygous genotype, the *APOE-ε4* allele was significantly associated with an earlier age at onset of disease regardless of ethnic group, but the effects of *APOE-ε4* heterozygosity on age-at-onset were limited to white, non-Hispanics. *APOE-ε4* showed similar effects in women and men.

The success of the NIA-LOAD FBS can be measured by the broad use of the data and samples (table 4). Over 67,000 biological samples collected as part of the NIA-LOAD FBS have been distributed to approved investigators. These samples have been analyzed, and the resulting data are available through the database of Genotypes and Phenotypes (dbGaP) and NIAGADS. In total, over 830 national and international investigators have requested data from the NIA-LOAD FBS, including the Alzheimer’s Disease Genetics Consortium (ADGC), the Alzheimer’s Disease Sequencing Project (ADSP), the Consortium for Alzheimer’s Disease Research (CADRE), the NIA-sponsored Alzheimer’s Disease Research Centers (ADRCs) and the National Alzheimer’s Coordinating Center (NACC). Investigators have generated 126 publications from the data and samples (Supplement 4).

Multiplex families are more difficult to recruit for research studies, sometimes taking several years to identify all of the affected relatives in a single family and complete their phenotyping. Large multiplex families are often thought to be too rare to be relevant. However, such families may actually be much more common than assumed, and there are families in isolated geographical regions where very large sibships occur.^38-40^ Families with multiple ill members can more easily be found and ascertained for such studies. With the increased use of virtual visits, the ability to longitudinally follow these families is becoming easier. These large multiplex families may hold the key to survival without dementia despite carrying a disease-causing variant.

The ultimate aims of genetic studies of complex diseases, such as AD, are to identify causal biological pathways that might provide targets for therapeutic development. However, diseases such as AD are multigenic and it is likely that there are multiple strategies to accomplish this difficult task. A significant proportion of the heritability remains unexplained but may be due to rare, inherited, moderately penetrant variants unique to individual families. The NIA-LOAD FBS remains a critical component of the overall strategy to tackle AD genetics in the United States that includes the Alzheimer’s Disease Genetics Consortium, the Alzheimer’s Disease Sequencing Project, the NCRAD and NIAGADS. Together these groups have created a large collaborative international effort to combine data worldwide in pursuit of uncovering the relevant genetic risk factors for AD with the hope that patterns may emerge that ultimately will uncover biologic pathways underlying the illness.

## Supporting information

NIA-LOAD FBS Supplemental Tables and Figures

## Data Availability

All clinical and genetic data are available to qualified investigators at the National Institute on Aging Genetics of Alzheimer's Disease Storage Site (NIAGADS). Biological Samples related to these data are available to qualified investigators from the National Centralized Repository for Alzheimer's Disease and Related Disorders (NCRAD)

https://www.niagads.org

https://ncrad.iu.edu

## Acknowledgements

The NIA-LOAD FBS supported the collection of samples used in this study through National Institute on Aging (NIA) grants U24AG026395, U24AG021886, R01AG041797 and U24AG056270. Additional families were contributed to the NIA-LOAD FBS through NIH grants: R01AG028786, R01AG027944, RO1AG027944, RF1AG054074, U01AG052410. We thank contributors, including the Alzheimer’s Disease Centers who collected samples used in this study, as well as patients and their families, whose help and participation made this work possible.

