## Supplementary material for "The National Institute on Aging Late Onset Alzheimer’s Disease Family Based Study: A Critical Component of the International Effort to Understand Alzheimer’s Disease": NIA-LOAD FBS Supplemental Tables and Figures: NIA-LOAD FBS.v1.Supplemental Tables and Figures .docx

| Supplemental Table 1. Samples and Genetic Data | | |
| --- | --- | --- |
| Samples | **Families** | **Individuals** |
| DNA * | 1,446 (82.3%) | 7,925 (82%) |
| PBMCs | 152 (8.7%) | 322 (3.3%) |
| Genetic Data | Families | Individuals |
| *APOE* genotyping | 1,260 (71.8%) | 7,601(78.5%) |
| GWAS | 1,340 (76.3%) | 7,012 (72.4%) |
| Whole Exome Sequencing | 926 (52.7%) | 1,650 (17%) |
| Whole Genome Sequencing | 414 (23.6%) | 1,501 (15.5%) |
| RNA sequencing and DNA methylation | 240 (16.6%) | 330 (50.4%) |

*Good quality DNA

| Supplemental Table 2. Ethnic distribution of families by affection status. | | | |
| --- | --- | --- | --- |
| Families | Affected (%) | Unaffected (%) | Total |
| African American | 142 (4.1%) | 175 (3.5%) | 317 (3.7%) |
| Caribbean Hispanic | 1,284 (36.7%) | 1,799 (35.9%) | 3,083 (36.3%) |
| Other | 64 (1.8%) | 98 (1.9%) | 162 (1.9%) |
| White, Non-Hispanic | 2,006 (57.4%) | 2,935 (58.6%) | 4,941 (58.1%) |
| Total | 3,496 | 5,007 | 8,503 |

**Supplemental figure 1. NIA-LOAD Family Based Sites in the United States.**

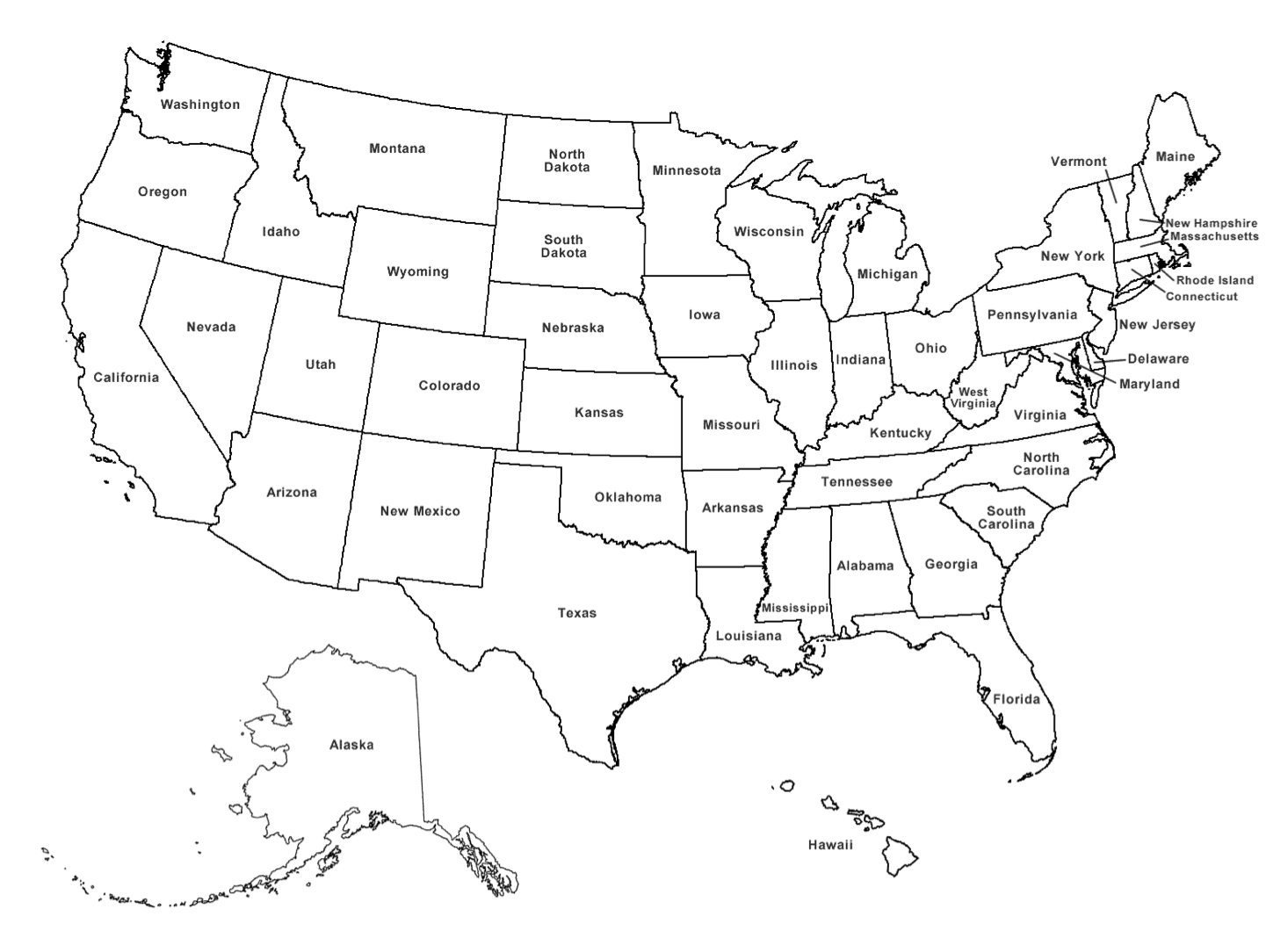

CWR

UP

IU

UM

UTSW

UW

Mayo

Rush

CU

WUStL

Site abbreviations: CU-Columbia University Medical Center, UM-University of Miami, UP-University of Pittsburgh, IU-Indiana University, WUStL-Washington University St Louis, Rush-Rush University Medical Center, Mayo-Mayo Clinic, Rochester, UTSW-University of Texas Southwestern Medical Center, UW-University of Washington.

Supplemental Figures 2a and 2b. The upper figure (2a) shows the effect of APOE genotype on that age-at-onset of AD in women, and the lower figure (2b) shows the effects of APOE in men.

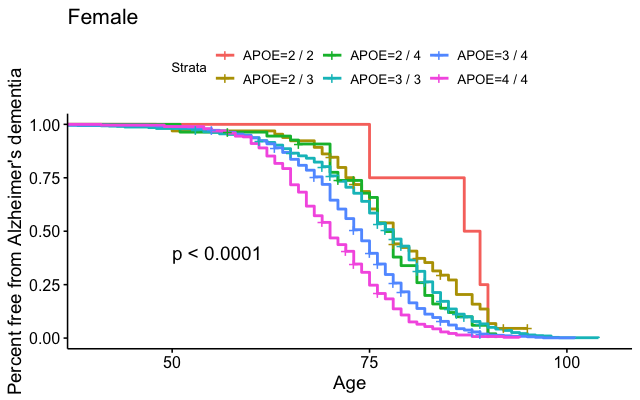

women

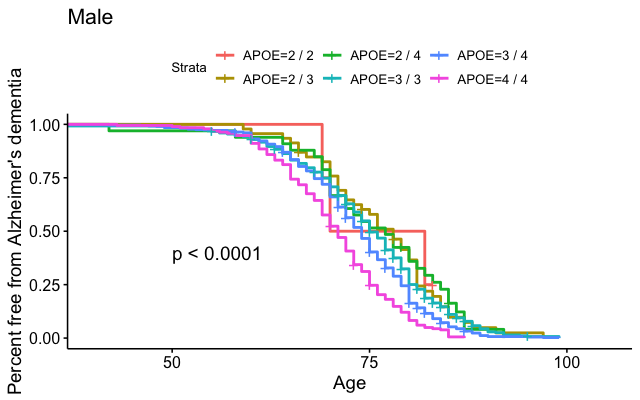

men

Supplemental Figures 3a-d. 3a shows the overall effects of *APOE* genotype in the entire cohort, 3b is restricted to white, non-Hispanics, 3c is restricted to Hispanics and 3d to African Americans.

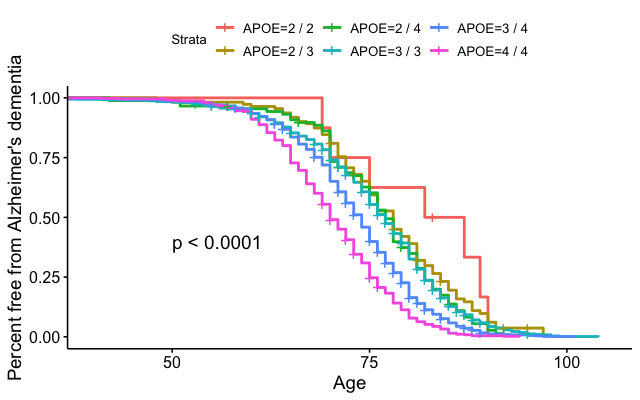

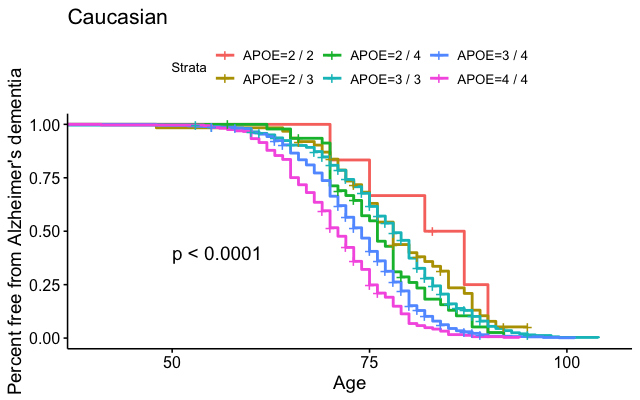

White, non-Hispanic

Overall

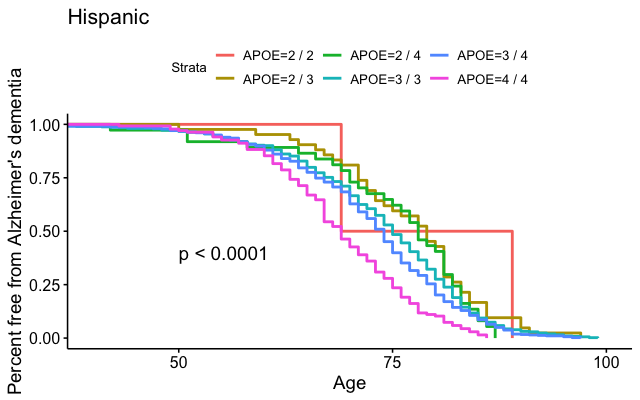

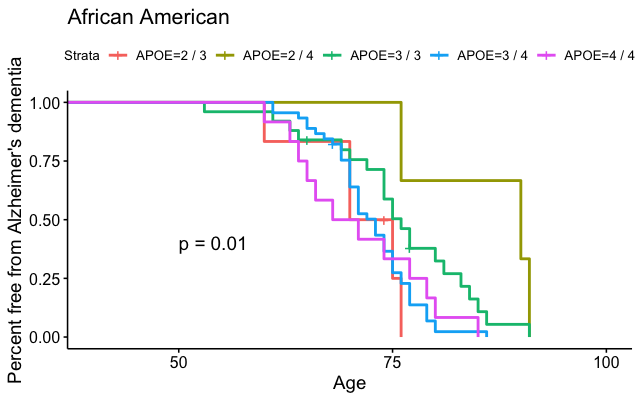

African American

Caribbean Hispanic

**Supplement 4. Manuscripts using Data or Samples from the NIA-LOAD Family Based Study**

1. Allen M, Zou F, Chai HS, Younkin CS, Crook J, Pankratz VS, Carrasquillo MM, Rowley CN, Nair AA, Middha S, Maharjan S, Nguyen T, Ma L, Malphrus KG, Palusak R, Lincoln S, Bisceglio G, Georgescu C, Schultz D, Rakhshan F, Kolbert CP, Jen J, Haines JL, Mayeux R, Pericak-Vance MA, Farrer LA, Schellenberg GD, Petersen RC, Graff-Radford NR, Dickson DW, Younkin SG, Ertekin-Taner N; Alzheimer's Disease Genetics Consortium (ADGC), Apostolova LG, Arnold SE, Baldwin CT, Barber R, Barmada MM, Beach T, Beecham GW, Beekly D, Bennett DA, Bigio EH, Bird TD, Blacker D, Boeve BF, Bowen JD, Boxer A, Burke JR, Buros J, Buxbaum JD, Cairns NJ, Cantwell LB, Cao C, Carlson CS, Carney RM, Carroll SL, Chui HC, Clark DG, Corneveaux J, Cotman CW, Crane PK, Cruchaga C, Cummings JL, De Jager PL, DeCarli C, DeKosky ST, Demirci FY, Diaz-Arrastia R, Dick M, Dombroski BA, Duara R, Ellis WD, Evans D, Faber KM, Fallon KB, Farlow MR, Ferris S, Foroud TM, Frosch M, Galasko DR, Gallins PJ, Ganguli M, Gearing M, Geschwind DH, Ghetti B, Gilbert JR, Gilman S, Giordani B, Glass JD, Goate AM, Green RC, Growdon JH, Hakonarson H, Hamilton RL, Hardy J, Harrell LE, Head E, Honig LS, Huentelman MJ, Hulette CM, Hyman BT, Jarvik GP, Jicha GA, Jin LW, Jun G, Kamboh MI, Karlawish J, Karydas A, Kauwe JS, Kaye JA, Kennedy N, Kim R, Koo EH, Kowall NW, Kramer P, Kukull WA, Lah JJ, Larson EB, Levey AI, Lieberman AP, Lopez OL, Lunetta KL, Mack WJ, Marson DC, Martin ER, Martiniuk F, Mash DC, Masliah E, McCormick WC, McCurry SM, McDavid AN, McKee AC, Mesulam M, Miller BL, Miller CA, Miller JW, Montine TJ, Morris JC, Myers AJ, Naj AC, Nowotny P, Parisi JE, Perl DP, Peskind E, Poon WW, Potter H, Quinn JF, Raj A, Rajbhandary RA, Raskind M, Reiman EM, Reisberg B, Reitz C, Ringman JM, Roberson ED, Rogaeva E, Rosenberg RN, Sano M, Saykin AJ, Schneider JA, Schneider LS, Seeley W, Shelanski ML, Slifer MA, Smith CD, Sonnen JA, Spina S, St George-Hyslop P, Stern RA, Tanzi RE, Trojanowski JQ, Troncoso JC, Tsuang DW, Van Deerlin VM, Vardarajan BN, Vinters HV, Vonsattel JP, Wang LS, Weintraub S, Welsh-Bohmer KA, Williamson J, Woltjer RL. Novel late-onset Alzheimer disease loci variants associate with brain gene expression. Neurology. 2012 Jul 17;79(3):221-8. PubMed PMID: 22722634; PubMed Central PMCID: PMC3398432.
2. Andrews SJ, Fulton-Howard B, O'Reilly P, Marcora E, Goate AM; collaborators of the Alzheimer's Disease Genetics Consortium. Causal Associations Between Modifiable Risk Factors and the Alzheimer's Phenome. Ann Neurol. 2021 Jan;89(1):54-65. PMID: 32996171.
3. Baker E, Sims R, Leonenko G, Frizzati A, Harwood JC, Grozeva D; GERAD/PERADES; CHARGE; ADGC; EADI; IGAP consortia, Morgan K, Passmore P, Holmes C, Powell J, Brayne C, Gill M, Mead S, Bossù P, Spalletta G, Goate AM, Cruchaga C, Maier W, Heun R, Jessen F, Peters O, Dichgans M, FröLich L, Ramirez A, Jones L, Hardy J, Ivanov D, Hill M, Holmans P, Allen ND, Morgan BP, Seshadri S, Schellenberg GD, Amouyel P, Williams J, Escott-Price V. Gene-based analysis in HRC imputed genome wide association data identifies three novel genes for Alzheimer's disease. PLoS One. 2019 Jul 8;14(7):e0218111. PMID: 31283791
4. Barral S, Bird T, Goate A, Farlow MR, Diaz-Arrastia R, Bennett DA, Graff-Radford N, Boeve BF, Sweet RA, Stern Y, Wilson RS, Foroud T, Ott J, Mayeux R; National Institute on Aging Late-Onset Alzheimer's Disease Genetics Study. Genotype patterns at PICALM, CR1, BIN1, CLU, and APOE genes are associated with episodic memory. Neurology. 2012 May 8;78(19):1464-71. PubMed PMID: 22539578; PubMed Central PMCID: PMC3345618.
5. Barral S, Cheng R, Reitz C, Vardarajan B, Lee J, Kunkle B, Beecham G, Cantwell LS, Pericak-Vance MA, Farrer LA, Haines JL, Goate AM, Foroud T, Boerwinkle E, Schellenberg GD, Mayeux R. Linkage analyses in Caribbean Hispanic families identify novel loci associated with familial late-onset Alzheimer's disease. Alzheimers Dement. 2015 Dec;11(12):1397-1406. PubMed PMID: 26433351; PubMed Central PMCID: PMC4690771.
6. Barral S, Vardarajan BN, Reyes-Dumeyer D, Faber KM, Bird TD, Tsuang D, Bennett DA, Rosenberg R, Boeve BF, Graff-Radford NR, Goate AM, Farlow M, Lantigua R, Medrano MZ, Wang X, Kamboh MI, Barmada MM, Schaid DJ, Foroud TM, Weamer EA, Ottman R, Sweet RA, Mayeux R; NIA-LOAD/NCRAD Family Study Group. Genetic variants associated with susceptibility to psychosis in late-onset Alzheimer's disease families. Neurobiol Aging. 2015 Nov;36(11):3116. e9-3116.e16. PubMed PMID: 26359528; PubMed Central PMCID: PMC4609604.
7. Beecham GW, Hamilton K, Naj AC, Martin ER, Huentelman M, Myers AJ, Corneveaux JJ, Hardy J, Vonsattel JP, Younkin SG, Bennett DA, De Jager PL, Larson EB, Crane PK, Kamboh MI, Kofler JK, Mash DC, Duque L, Gilbert JR, Gwirtsman H, Buxbaum JD, Kramer P, Dickson DW, Farrer LA, Frosch MP, Ghetti B, Haines JL, Hyman BT, Kukull WA, Mayeux RP, Pericak-Vance MA, Schneider JA, Trojanowski JQ, Reiman EM; Alzheimer's Disease Genetics Consortium (ADGC), Schellenberg GD, Montine TJ. Genome-wide association meta-analysis of neuropathologic features of Alzheimer's disease and related dementias. PLoS Genet. 2014 Sep 4;10(9):e1004606. Sep. Erratum in: PLoS Genet. 2014 Nov;10(11):e1004867. PubMed PMID: 25188341; PubMed Central PMCID: PMC4154667.
8. Beecham GW, Vardarajan B, Blue E, Bush W, Jaworski J, Barral S, DeStefano A, Hamilton-Nelson K, Kunkle B, Martin ER, Naj A, Rajabli F, Reitz C, Thornton T, van Duijn C, Goate A, Seshadri S, Farrer LA, Boerwinkle E, Schellenberg G, Haines JL, Wijsman E, Mayeux R, Pericak-Vance MA; Alzheimer's Disease Sequencing Project. Rare genetic variation implicated in non-Hispanic white families with Alzheimer disease. Neurol Genet. 2018 Nov 21;4(6):e286. PubMed PMID: 30569016; PubMed Central PMCID: PMC6278241.
9. Benitez BA, Jin SC, Guerreiro R, Graham R, Lord J, Harold D, Sims R, Lambert JC, Gibbs JR, Bras J, Sassi C, Harari O, Bertelsen S, Lupton MK, Powell J, Bellenguez C, Brown K, Medway C, Haddick PC, van der Brug MP, Bhangale T, Ortmann W, Behrens T, Mayeux R, Pericak-Vance MA, Farrer LA, Schellenberg GD, Haines JL, Turton J, Braae A, Barber I, Fagan AM, Holtzman DM, Morris JC; 3C Study Group; EADI consortium; Alzheimer's Disease Genetic Consortium (ADGC); Alzheimer's Disease Neuroimaging Initiative (ADNI); GERAD Consortium, Williams J, Kauwe JS, Amouyel P, Morgan K, Singleton A, Hardy J, Goate AM, Cruchaga C. Missense variant in TREML2 protects against Alzheimer's disease. Neurobiol Aging. 2014 Jun;35(6):1510.e19-26. PubMed PMID: 24439484; PubMed Central PMCID: PMC3961557.
10. Bis JC, Jian X, Kunkle BW, Chen Y, Hamilton-Nelson KL, Bush WS, Salerno WJ, Lancour D, Ma Y, Renton AE, Marcora E, Farrell JJ, Zhao Y, Qu L, Ahmad S, Amin N, Amouyel P, Beecham GW, Below JE, Campion D, Charbonnier C, Chung J, Crane PK, Cruchaga C, Cupples LA, Dartigues JF, Debette S, Deleuze JF, Fulton L, Gabriel SB, Genin E, Gibbs RA, Goate A, Grenier-Boley B, Gupta N, Haines JL, Havulinna AS, Helisalmi S, Hiltunen M, Howrigan DP, Ikram MA, Kaprio J, Konrad J, Kuzma A, Lander ES, Lathrop M, Lehtimäki T, Lin H, Mattila K, Mayeux R, Muzny DM, Nasser W, Neale B, Nho K, Nicolas G, Patel D, Pericak-Vance MA, Perola M, Psaty BM, Quenez O, Rajabli F, Redon R, Reitz C, Remes AM, Salomaa V, Sarnowski C, Schmidt H, Schmidt M, Schmidt R, Soininen H, Thornton TA, Tosto G, Tzourio C, van der Lee SJ, van Duijn CM, Vardarajan B, Wang W, Wijsman E, Wilson RK, Witten D, Worley KC, Zhang X; Alzheimer’s Disease Sequencing Project, Bellenguez C, Lambert JC, Kurki MI, Palotie A, Daly M, Boerwinkle E, Lunetta KL, Destefano AL, Dupuis J, Martin ER, Schellenberg GD, Seshadri S, Naj AC, Fornage M, Farrer LA. Whole exome sequencing study identifies novel rare and common Alzheimer's-Associated variants involved in immune response and transcriptional regulation. Mol Psychiatry. 2018 Aug 14. doi: 10.1038/s41380-018-0112-7. [PubMed PMID: 30108311; PubMed Central PMCID: PMC6375806.
11. Blue EE, Bis JC, Dorschner MO, Tsuang DW, Barral SM, Beecham G, Below JE, Bush WS, Butkiewicz M, Cruchaga C, DeStefano A, Farrer LA, Goate A, Haines J, Jaworski J, Jun G, Kunkle B, Kuzma A, Lee JJ, Lunetta KL, Ma Y, Martin E, Naj A, Nato AQ, Navas P, Nguyen H, Reitz C, Reyes D, Salerno W, Schellenberg GD, Seshadri S, Sohi H, Thornton TA, Valadares O, van Duijn C, Vardarajan BN, Wang LS, Boerwinkle E, Dupuis J, Pericak-Vance MA, Mayeux R, Wijsman EM; on behalf of the Alzheimer’s Disease Sequencing Project. Genetic Variation in Genes Underlying Diverse Dementias May Explain a Small Proportion of Cases in the Alzheimer's Disease Sequencing Project. Dement Geriatr Cogn Disord. 2018;45(1-2):1-17. PubMed PMID: 29486463; PubMed Central PMCID: PMC5971141.
12. Brainstorm Consortium, Anttila V, Bulik-Sullivan B, Finucane HK, Walters RK, Bras J, Duncan L, Escott-Price V, Falcone GJ, Gormley P, Malik R, Patsopoulos NA, Ripke S, Wei Z, Yu D, Lee PH, Turley P, Grenier-Boley B, Chouraki V, Kamatani Y, Berr C, Letenneur L, Hannequin D, Amouyel P, Boland A, Deleuze JF, Duron E, Vardarajan BN, Reitz C, Goate AM, Huentelman MJ, Kamboh MI, Larson EB, Rogaeva E, St George-Hyslop P, Hakonarson H, Kukull WA, Farrer LA, Barnes LL, Beach TG, Demirci FY, Head E, Hulette CM, Jicha GA, Kauwe JSK, Kaye JA, Leverenz JB, Levey AI, Lieberman AP, Pankratz VS, Poon WW, Quinn JF, Saykin AJ, Schneider LS, Smith AG, Sonnen JA, Stern RA, Van Deerlin VM, Van Eldik LJ, Harold D, Russo G, Rubinsztein DC, Bayer A, Tsolaki M, Proitsi P, Fox NC, Hampel H, Owen MJ, Mead S, Passmore P, Morgan K, Nöthen MM, Rossor M, Lupton MK, Hoffmann P, Kornhuber J, Lawlor B, McQuillin A, Al-Chalabi A, Bis JC, Ruiz A, Boada M, Seshadri S, Beiser A, Rice K, van der Lee SJ, De Jager PL, Geschwind DH, Riemenschneider M, Riedel-Heller S, Rotter JI, Ransmayr G, Hyman BT, Cruchaga C, Alegret M, Winsvold B, Palta P, Farh KH, Cuenca-Leon E, Furlotte N, Kurth T, Ligthart L, Terwindt GM, Freilinger T, Ran C, Gordon SD, Borck G, Adams HHH, Lehtimäki T, Wedenoja J, Buring JE, Schürks M, Hrafnsdottir M, Hottenga JJ, Penninx B, Artto V, Kaunisto M, Vepsäläinen S, Martin NG, Montgomery GW, Kurki MI, Hämäläinen E, Huang H, Huang J, Sandor C, Webber C, Muller-Myhsok B, Schreiber S, Salomaa V, Loehrer E, Göbel H, Macaya A, Pozo-Rosich P, Hansen T, Werge T, Kaprio J, Metspalu A, Kubisch C, Ferrari MD, Belin AC, van den Maagdenberg AMJM, Zwart JA, Boomsma D, Eriksson N, Olesen J, Chasman DI, Nyholt DR, Avbersek A, Baum L, Berkovic S, Bradfield J, Buono RJ, Catarino CB, Cossette P, De Jonghe P, Depondt C, Dlugos D, Ferraro TN, French J, Hjalgrim H, Jamnadas-Khoda J, Kälviäinen R, Kunz WS, Lerche H, Leu C, Lindhout D, Lo W, Lowenstein D, McCormack M, Møller RS, Molloy A, Ng PW, Oliver K, Privitera M, Radtke R, Ruppert AK, Sander T, Schachter S, Schankin C, Scheffer I, Schoch S, Sisodiya SM, Smith P, Sperling M, Striano P, Surges R, Thomas GN, Visscher F, Whelan CD, Zara F, Heinzen EL, Marson A, Becker F, Stroink H, Zimprich F, Gasser T, Gibbs R, Heutink P, Martinez M, Morris HR, Sharma M, Ryten M, Mok KY, Pulit S, Bevan S, Holliday E, Attia J, Battey T, Boncoraglio G, Thijs V, Chen WM, Mitchell B, Rothwell P, Sharma P, Sudlow C, Vicente A, Markus H, Kourkoulis C, Pera J, Raffeld M, Silliman S, Boraska Perica V, Thornton LM, Huckins LM, William Rayner N, Lewis CM, Gratacos M, Rybakowski F, Keski-Rahkonen A, Raevuori A, Hudson JI, Reichborn-Kjennerud T, Monteleone P, Karwautz A, Mannik K, Baker JH, O'Toole JK, Trace SE, Davis OSP, Helder SG, Ehrlich S, Herpertz-Dahlmann B, Danner UN, van Elburg AA, Clementi M, Forzan M, Docampo E, Lissowska J, Hauser J, Tortorella A, Maj M, Gonidakis F, Tziouvas K, Papezova H, Yilmaz Z, Wagner G, Cohen-Woods S, Herms S, Julià A, Rabionet R, Dick DM, Ripatti S, Andreassen OA, Espeseth T, Lundervold AJ, Steen VM, Pinto D, Scherer SW, Aschauer H, Schosser A, Alfredsson L, Padyukov L, Halmi KA, Mitchell J, Strober M, Bergen AW, Kaye W, Szatkiewicz JP, Cormand B, Ramos-Quiroga JA, Sánchez-Mora C, Ribasés M, Casas M, Hervas A, Arranz MJ, Haavik J, Zayats T, Johansson S, Williams N, Dempfle A, Rothenberger A, Kuntsi J, Oades RD, Banaschewski T, Franke B, Buitelaar JK, Arias Vasquez A, Doyle AE, Reif A, Lesch KP, Freitag C, Rivero O, Palmason H, Romanos M, Langley K, Rietschel M, Witt SH, Dalsgaard S, Børglum AD, Waldman I, Wilmot B, Molly N, Bau CHD, Crosbie J, Schachar R, Loo SK, McGough JJ, Grevet EH, Medland SE, Robinson E, Weiss LA, Bacchelli E, Bailey A, Bal V, Battaglia A, Betancur C, Bolton P, Cantor R, Celestino-Soper P, Dawson G, De Rubeis S, Duque F, Green A, Klauck SM, Leboyer M, Levitt P, Maestrini E, Mane S, De-Luca DM, Parr J, Regan R, Reichenberg A, Sandin S, Vorstman J, Wassink T, Wijsman E, Cook E, Santangelo S, Delorme R, Rogé B, Magalhaes T, Arking D, Schulze TG, Thompson RC, Strohmaier J, Matthews K, Melle I, Morris D, Blackwood D, McIntosh A, Bergen SE, Schalling M, Jamain S, Maaser A, Fischer SB, Reinbold CS, Fullerton JM, Guzman-Parra J, Mayoral F, Schofield PR, Cichon S, Mühleisen TW, Degenhardt F, Schumacher J, Bauer M, Mitchell PB, Gershon ES, Rice J, Potash JB, Zandi PP, Craddock N, Ferrier IN, Alda M, Rouleau GA, Turecki G, Ophoff R, Pato C, Anjorin A, Stahl E, Leber M, Czerski PM, Cruceanu C, Jones IR, Posthuma D, Andlauer TFM, Forstner AJ, Streit F, Baune BT, Air T, Sinnamon G, Wray NR, MacIntyre DJ, Porteous D, Homuth G, Rivera M, Grove J, Middeldorp CM, Hickie I, Pergadia M, Mehta D, Smit JH, Jansen R, de Geus E, Dunn E, Li QS, Nauck M, Schoevers RA, Beekman AT, Knowles JA, Viktorin A, Arnold P, Barr CL, Bedoya-Berrio G, Bienvenu OJ, Brentani H, Burton C, Camarena B, Cappi C, Cath D, Cavallini M, Cusi D, Darrow S, Denys D, Derks EM, Dietrich A, Fernandez T, Figee M, Freimer N, Gerber G, Grados M, Greenberg E, Hanna GL, Hartmann A, Hirschtritt ME, Hoekstra PJ, Huang A, Huyser C, Illmann C, Jenike M, Kuperman S, Leventhal B, Lochner C, Lyon GJ, Macciardi F, Madruga-Garrido M, Malaty IA, Maras A, McGrath L, Miguel EC, Mir P, Nestadt G, Nicolini H, Okun MS, Pakstis A, Paschou P, Piacentini J, Pittenger C, Plessen K, Ramensky V, Ramos EM, Reus V, Richter MA, Riddle MA, Robertson MM, Roessner V, Rosário M, Samuels JF, Sandor P, Stein DJ, Tsetsos F, Van Nieuwerburgh F, Weatherall S, Wendland JR, Wolanczyk T, Worbe Y, Zai G, Goes FS, McLaughlin N, Nestadt PS, Grabe HJ, Depienne C, Konkashbaev A, Lanzagorta N, Valencia-Duarte A, Bramon E, Buccola N, Cahn W, Cairns M, Chong SA, Cohen D, Crespo-Facorro B, Crowley J, Davidson M, DeLisi L, Dinan T, Donohoe G, Drapeau E, Duan J, Haan L, Hougaard D, Karachanak-Yankova S, Khrunin A, Klovins J, Kučinskas V, Lee Chee Keong J, Limborska S, Loughland C, Lönnqvist J, Maher B, Mattheisen M, McDonald C, Murphy KC, Nenadic I, van Os J, Pantelis C, Pato M, Petryshen T, Quested D, Roussos P, Sanders AR, Schall U, Schwab SG, Sim K, So HC, Stögmann E, Subramaniam M, Toncheva D, Waddington J, Walters J, Weiser M, Cheng W, Cloninger R, Curtis D, Gejman PV, Henskens F, Mattingsdal M, Oh SY, Scott R, Webb B, Breen G, Churchhouse C, Bulik CM, Daly M, Dichgans M, Faraone SV, Guerreiro R, Holmans P, Kendler KS, Koeleman B, Mathews CA, Price A, Scharf J, Sklar P, Williams J, Wood NW, Cotsapas C, Palotie A, Smoller JW, Sullivan P, Rosand J, Corvin A, Neale BM, Schott JM, Anney R, Elia J, Grigoroiu-Serbanescu M, Edenberg HJ, Murray R. Analysis of shared heritability in common disorders of the brain. Science. 2018 Jun 22;360(6395). PubMed PMID: 29930110; PubMed Central PMCID: PMC6097237.
13. Broce IJ, Tan CH, Fan CC, Jansen I, Savage JE, Witoelar A, Wen N, Hess CP, Dillon WP, Glastonbury CM, Glymour M, Yokoyama JS, Elahi FM, Rabinovici GD, Miller BL, Mormino EC, Sperling RA, Bennett DA, McEvoy LK, Brewer JB, Feldman HH, Hyman BT, Pericak-Vance M, Haines JL, Farrer LA, Mayeux R, Schellenberg GD, Yaffe K, Sugrue LP, Dale AM, Posthuma D, Andreassen OA, Karch CM, Desikan RS. Dissecting the genetic relationship between cardiovascular risk factors and Alzheimer's disease. Acta Neuropathol. 2019 Feb;137(2):209-226. PubMed PMID: 30413934; PubMed Central PMCID: PMC6358498.
14. Chauhan G, Adams HHH, Satizabal CL, Bis JC, Teumer A, Sargurupremraj M, Hofer E, Trompet S, Hilal S, Smith AV, Jian X, Malik R, Traylor M, Pulit SL, Amouyel P, Mazoyer B, Zhu YC, Kaffashian S, Schilling S, Beecham GW, Montine TJ, Schellenberg GD, Kjartansson O, Guðnason V, Knopman DS, Griswold ME, Windham BG, Gottesman RF, Mosley TH, Schmidt R, Saba Y, Schmidt H, Takeuchi F, Yamaguchi S, Nabika T, Kato N, Rajan KB, Aggarwal NT, De Jager PL, Evans DA, Psaty BM, Rotter JI, Rice K, Lopez OL, Liao J, Chen C, Cheng CY, Wong TY, Ikram MK, van der Lee SJ, Amin N, Chouraki V, DeStefano AL, Aparicio HJ, Romero JR, Maillard P, DeCarli C, Wardlaw JM, Hernández MDCV, Luciano M, Liewald D, Deary IJ, Starr JM, Bastin ME, Muñoz Maniega S, Slagboom PE, Beekman M, Deelen J, Uh HW, Lemmens R, Brodaty H, Wright MJ, Ames D, Boncoraglio GB, Hopewell JC, Beecham AH, Blanton SH, Wright CB, Sacco RL, Wen W, Thalamuthu A, Armstrong NJ, Chong E, Schofield PR, Kwok JB, van der Grond J, Stott DJ, Ford I, Jukema JW, Vernooij MW, Hofman A, Uitterlinden AG, van der Lugt A, Wittfeld K, Grabe HJ, Hosten N, von Sarnowski B, Völker U, Levi C, Jimenez-Conde J, Sharma P, Sudlow CLM, Rosand J, Woo D, Cole JW, Meschia JF, Slowik A, Thijs V, Lindgren A, Melander O, Grewal RP, Rundek T, Rexrode K, Rothwell PM, Arnett DK, Jern C, Johnson JA, Benavente OR, Wasssertheil-Smoller S, Lee JM, Wong Q, Mitchell BD, Rich SS, McArdle PF, Geerlings MI, van der Graaf Y, de Bakker PIW, Asselbergs FW, Srikanth V, Thomson R, McWhirter R, Moran C, Callisaya M, Phan T, Rutten-Jacobs LCA, Bevan S, Tzourio C, Mather KA, Sachdev PS, van Duijn CM, Worrall BB, Dichgans M, Kittner SJ, Markus HS, Ikram MA, Fornage M, Launer LJ, Seshadri S, Longstreth WT Jr, Debette S; Stroke Genetics Network (SiGN), the International Stroke Genetics Consortium (ISGC), METASTROKE, Alzheimer's Disease Genetics Consortium (ADGC), and the Neurology Working Group of the Cohorts for Heart and Aging Research in Genomic Epidemiology (CHARGE) Consortium. Genetic and lifestyle risk factors for MRI-defined brain infarcts in a population-based setting. Neurology. 2019 92(5):e486-503. PubMed PMID: 30651383; PubMed Central PMCID: PMC6369905.
15. Choi KY, Lee JJ, Gunasekaran TI, Kang S, Lee W, Jeong J, Lim HJ, Zhang X, Zhu C, Won SY, Choi YY, Seo EH, Lee SC, Gim J, Chung JY, Chong A, Byun MS, Seo S, Ko PW, Han JW, McLean C, Farrell J, Lunetta KL, Miyashita A, Hara N, Won S, Choi SM, Ha JM, Jeong JH, Kuwano R, Song MK, An SSA, Lee YM, Park KW, Lee HW, Choi SH, Rhee S, Song WK, Lee JS, Mayeux R, Haines JL, Pericak-Vance MA, Choo ILH, Nho K, Kim KW, Lee DY, Kim S, Kim BC, Kim H, Jun GR, Schellenberg GD, Ikeuchi T, Farrer LA, Lee KH, Neuroimaging Initative AD. *APOE* Promoter Polymorphism-219T/G is an Effect Modifier of the Influence of *APOE* ε4 on Alzheimer's Disease Risk in a Multiracial Sample. J Clin Med. 2019 Aug 16;8(8):1236. PMID: 31426376; PMCID: PMC6723529.
16. Chung J, Zhang X, Allen M, Wang X, Ma Y, Beecham G, Montine TJ, Younkin SG, Dickson DW, Golde TE, Price ND, Ertekin-Taner N, Lunetta KL, Mez J; Alzheimer’s Disease Genetics Consortium, Mayeux R, Haines JL, Pericak-Vance MA, Schellenberg G, Jun GR, Farrer LA. Genome-wide pleiotropy analysis of neuropathological traits related to Alzheimer's disease. Alzheimers Res Ther. 2018 Feb 20;10(1):22. PubMed PMID: 29458411; PubMed Central PMCID: PMC5819208.
17. Coppola G, Chinnathambi S, Lee JJ, Dombroski BA, Baker MC, Soto-Ortolaza AI, Lee SE, Klein E, Huang AY, Sears R, Lane JR, Karydas AM, Kenet RO, Biernat J, Wang LS, Cotman CW, Decarli CS, Levey AI, Ringman JM, Mendez MF, Chui HC, Le Ber I, Brice A, Lupton MK, Preza E, Lovestone S, Powell J, Graff-Radford N, Petersen RC, Boeve BF, Lippa CF, Bigio EH, Mackenzie I, Finger E, Kertesz A, Caselli RJ, Gearing M, Juncos JL, Ghetti B, Spina S, Bordelon YM, Tourtellotte WW, Frosch MP, Vonsattel JP, Zarow C, Beach TG, Albin RL, Lieberman AP, Lee VM, Trojanowski JQ, Van Deerlin VM, Bird TD, Galasko DR, Masliah E, White CL, Troncoso JC, Hannequin D, Boxer AL, Geschwind MD, Kumar S, Mandelkow EM, Wszolek ZK, Uitti RJ, Dickson DW, Haines JL, Mayeux R, Pericak-Vance MA, Farrer LA; Alzheimer's Disease Genetics Consortium, Ross OA, Rademakers R, Schellenberg GD, Miller BL, Mandelkow E, Geschwind DH. Evidence for a role of the rare p.A152T variant in MAPT in increasing the risk for FTD-spectrum and Alzheimer's diseases. Hum Mol Genet. 2012 Aug 1;21(15):3500-12. PubMed PMID: 22556362; PubMed Central PMCID: PMC3392107.
18. Cruchaga C, Del-Aguila JL, Saef B, Black K, Fernandez MV, Budde J, Ibanez L, Deming Y, Kapoor M, Tosto G, Mayeux RP, Holtzman DM, Fagan AM, Morris JC, Bateman RJ, Goate AM; Dominantly Inherited Alzheimer Network (DIAN); Disease Neuroimaging Initiative (ADNI); NIA-LOAD family study, Harari O. Polygenic risk score of sporadic late-onset Alzheimer's disease reveals a shared architecture with the familial and early-onset forms. Alzheimers Dement. 2018 Feb;14(2):205-214. PubMed PMID: 28943286; PubMed Central PMCID: PMC5803427.
19. Cruchaga C, Haller G, Chakraverty S, Mayo K, Vallania FL, Mitra RD, Faber K, Williamson J, Bird T, Diaz-Arrastia R, Foroud TM, Boeve BF, Graff-Radford NR, St Jean P, Lawson M, Ehm MG, Mayeux R, Goate AM; NIA-LOAD/NCRAD Family Study Consortium. Rare variants in APP, PSEN1 and PSEN2 increase risk for AD in late-onset Alzheimer's disease families. PLoS One. 2012;7(2):e31039. doi: 10.1371/journal.pone.0031039. Erratum in: PLoS One. 2012;7(5): doi/10.1371/annotation/c92e16da-7733-421d-b063-1db19488daa6. Haller, Gabe [added].. PubMed PMID: 22312439; PubMed Central PMCID: PMC3270040.
20. Cruchaga C, Kauwe JS, Harari O, Jin SC, Cai Y, Karch CM, Benitez BA, Jeng AT, Skorupa T, Carrell D, Bertelsen S, Bailey M, McKean D, Shulman JM, De Jager PL, Chibnik L, Bennett DA, Arnold SE, Harold D, Sims R, Gerrish A, Williams J, Van Deerlin VM, Lee VM, Shaw LM, Trojanowski JQ, Haines JL, Mayeux R, Pericak-Vance MA, Farrer LA, Schellenberg GD, Peskind ER, Galasko D, Fagan AM, Holtzman DM, Morris JC; GERAD Consortium; Alzheimer’s Disease Neuroimaging Initiative (ADNI); Alzheimer Disease Genetic Consortium (ADGC), Goate AM. GWAS of cerebrospinal fluid tau levels identifies risk variants for Alzheimer's disease. Neuron. 2013 Apr 24;78(2):256-68. PubMed PMID: 23562540; PubMed Central PMCID: PMC3664945.
21. Cukier HN, Kunkle BW, Vardarajan BN, Rolati S, Hamilton-Nelson KL, Kohli MA, Whitehead PL, Dombroski BA, Van Booven D, Lang R, Dykxhoorn DM, Farrer LA, Cuccaro ML, Vance JM, Gilbert JR, Beecham GW, Martin ER, Carney RM, Mayeux R, Schellenberg GD, Byrd GS, Haines JL, Pericak-Vance MA; Alzheimer's Disease Genetics Consortium. ABCA7 frameshift deletion associated with Alzheimer disease in African Americans. Neurol Genet. 2016 May 17;2(3):e79. PubMed PMID: 27231719; PubMed Central PMCID: PMC4871806.
22. DeMichele-Sweet MAA, Weamer EA, Klei L, Vrana DT, Hollingshead DJ, Seltman HJ, Sims R, Foroud T, Hernandez I, Moreno-Grau S, Tárraga L, Boada M, Ruiz A, Williams J, Mayeux R, Lopez OL, Sibille EL, Kamboh MI, Devlin B, Sweet RA. Genetic risk of schizophrenia and psychosis in Alzheimer’s disease. Mol Psychiatry. 2018 Apr;23(4):963-972. PMID: 28461698
23. Deming Y, Dumitrescu L, Barnes LL, Thambisetty M, Kunkle B, Gifford KA, Bush WS, Chibnik LB, Mukherjee S, De Jager PL, Kukull W, Huentelman M, Crane PK, Resnick SM, Keene CD, Montine TJ, Schellenberg GD, Haines JL, Zetterberg H, Blennow K, Larson EB, Johnson SC, Albert M, Moghekar A, Del Aguila JL, Fernandez MV, Budde J, Hassenstab J, Fagan AM, Riemenschneider M, Petersen RC, Minthon L, Chao MJ, Van Deerlin VM, Lee VM, Shaw LM, Trojanowski JQ, Peskind ER, Li G, Davis LK, Sealock JM, Cox NJ; Alzheimer’s Disease Neuroimaging Initiative (ADNI); Alzheimer Disease Genetics Consortium (ADGC), Goate AM, Bennett DA, Schneider JA, Jefferson AL, Cruchaga C, Hohman TJ. Sex-specific genetic predictors of Alzheimer's disease biomarkers. Acta Neuropathol. 2018 Dec;136(6):857-872. PubMed PMID: 29967939; PubMed Central PMCID: PMC6280657.
24. Deming Y, Li Z, Kapoor M, Harari O, Del-Aguila JL, Black K, Carrell D, Cai Y, Fernandez MV, Budde J, Ma S, Saef B, Howells B, Huang KL, Bertelsen S, Fagan AM, Holtzman DM, Morris JC, Kim S, Saykin AJ, De Jager PL, Albert M, Moghekar A, O'Brien R, Riemenschneider M, Petersen RC, Blennow K, Zetterberg H, Minthon L, Van Deerlin VM, Lee VM, Shaw LM, Trojanowski JQ, Schellenberg G, Haines JL, Mayeux R, Pericak-Vance MA, Farrer LA, Peskind ER, Li G, Di Narzo AF; Alzheimer’s Disease Neuroimaging Initiative (ADNI); Alzheimer Disease Genetic Consortium (ADGC), Kauwe JS, Goate AM, Cruchaga C. Genome-wide association study identifies four novel loci associated with Alzheimer's endophenotypes and disease modifiers. Acta Neuropathol. 2017 May;133(5):839-856. PubMed PMID: 28247064; PubMed Central PMCID: PMC5613285.
25. Desikan RS, Fan CC, Wang Y, Schork AJ, Cabral HJ, Cupples LA, Thompson WK, Besser L, Kukull WA, Holland D, Chen CH, Brewer JB, Karow DS, Kauppi K, Witoelar A, Karch CM, Bonham LW, Yokoyama JS, Rosen HJ, Miller BL, Dillon WP, Wilson DM, Hess CP, Pericak-Vance M, Haines JL, Farrer LA, Mayeux R, Hardy J, Goate AM, Hyman BT, Schellenberg GD, McEvoy LK, Andreassen OA, Dale AM. Genetic assessment of age-associated Alzheimer disease risk: Development and validation of a polygenic hazard score. PLoS Med. 2017 Mar 21;14(3):e1002258. doi: 10.1371/journal.pmed.1002258. eCollection 2017 Mar. Erratum in: PLoS Med. 2017 Mar 28;14 (3):e1002289. PubMed PMID: 28323831; PubMed Central PMCID: PMC5360219.
26. Desikan RS, Schork AJ, Wang Y, Thompson WK, Dehghan A, Ridker PM, Chasman DI, McEvoy LK, Holland D, Chen CH, Karow DS, Brewer JB, Hess CP, Williams J, Sims R, O'Donovan MC, Choi SH, Bis JC, Ikram MA, Gudnason V, DeStefano AL, van der Lee SJ, Psaty BM, van Duijn CM, Launer L, Seshadri S, Pericak-Vance MA, Mayeux R, Haines JL, Farrer LA, Hardy J, Ulstein ID, Aarsland D, Fladby T, White LR, Sando SB, Rongve A, Witoelar A, Djurovic S, Hyman BT, Snaedal J, Steinberg S, Stefansson H, Stefansson K, Schellenberg GD, Andreassen OA, Dale AM; Inflammation working group, IGAP and DemGene Investigators. Polygenic Overlap Between C-Reactive Protein, Plasma Lipids, and Alzheimer Disease. Circulation. 2015 Jun 9;131(23):2061-2069. doi: 10.1161/CIRCULATIONAHA.115.015489. PubMed PMID: 25862742; PubMed Central PMCID: PMC4677995.
27. Dumitrescu L, Barnes LL, Thambisetty M, Beecham G, Kunkle B, Bush WS, Gifford KA, Chibnik LB, Mukherjee S, De Jager PL, Kukull W, Crane PK, Resnick SM, Keene CD, Montine TJ, Schellenberg GD, Deming Y, Chao MJ, Huentelman M, Martin ER, Hamilton-Nelson K, Shaw LM, Trojanowski JQ, Peskind ER, Cruchaga C, Pericak-Vance MA, Goate AM, Cox NJ, Haines JL, Zetterberg H, Blennow K, Larson EB, Johnson SC, Albert M; Alzheimer’s Disease Genetics Consortium and the Alzheimer’s Disease Neuroimaging Initiative, Bennett DA, Schneider JA, Jefferson AL, Hohman TJ. Sex differences in the genetic predictors of Alzheimer's pathology. Brain. 2019 Sep 1;142(9):2581-2589. PMID: 31497858
28. Dumitrescu L, Mahoney ER, Mukherjee S, Lee ML, Bush WS, Engelman CD, Lu Q, Fardo DW, Trittschuh EH, Mez J, Kaczorowski C, Hernandez Saucedo H, Widaman KF, Buckley R, Properzi M, Mormino E, Yang HS, Harrison T, Hedden T, Nho K, Andrews SJ, Tommet D, Hadad N, Sanders RE, Ruderfer DM, Gifford KA, Moore AM, Cambronero F, Zhong X, Raghavan NS, Vardarajan B; Alzheimer’s Disease Neuroimaging Initiative (ADNI); Alzheimer’s Disease Genetics Consortium (ADGC), A4 Study Team, Pericak-Vance MA, Farrer LA, Wang LS, Cruchaga C, Schellenberg G, Cox NJ, Haines JL, Dirk Keene C, Saykin AJ, Larson EB, Sperling RA, Mayeux R, Bennett DA, Schneider JA, Crane PK, Jefferson AL, Hohman TJ. Genetic variants and functional pathways associated with resilience to Alzheimer's disease. Brain. 2020 Aug 1;143(8):2561-2575. doi: 10.1093/brain/awaa209. PMID: 32844198; PMCID: PMC7447518.
29. Escott-Price V, Bellenguez C, Wang LS, Choi SH, Harold D, Jones L, Holmans P, Gerrish A, Vedernikov A, Richards A, DeStefano AL, Lambert JC, Ibrahim-Verbaas CA, Naj AC, Sims R, Jun G, Bis JC, Beecham GW, Grenier-Boley B, Russo G, Thornton-Wells TA, Denning N, Smith AV, Chouraki V, Thomas C, Ikram MA, Zelenika D, Vardarajan BN, Kamatani Y, Lin CF, Schmidt H, Kunkle B, Dunstan ML, Vronskaya M; United Kingdom Brain Expression Consortium, Johnson AD, Ruiz A, Bihoreau MT, Reitz C, Pasquier F, Hollingworth P, Hanon O, Fitzpatrick AL, Buxbaum JD, Campion D, Crane PK, Baldwin C, Becker T, Gudnason V, Cruchaga C, Craig D, Amin N, Berr C, Lopez OL, De Jager PL, Deramecourt V, Johnston JA, Evans D, Lovestone S, Letenneur L, Hernández I, Rubinsztein DC, Eiriksdottir G, Sleegers K, Goate AM, Fiévet N, Huentelman MJ, Gill M, Brown K, Kamboh MI, Keller L, Barberger-Gateau P, McGuinness B, Larson EB, Myers AJ, Dufouil C, Todd S, Wallon D, Love S, Rogaeva E, Gallacher J, George-Hyslop PS, Clarimon J, Lleo A, Bayer A, Tsuang DW, Yu L, Tsolaki M, Bossù P, Spalletta G, Proitsi P, Collinge J, Sorbi S, Garcia FS, Fox NC, Hardy J, Naranjo MC, Bosco P, Clarke R, Brayne C, Galimberti D, Scarpini E, Bonuccelli U, Mancuso M, Siciliano G, Moebus S, Mecocci P, Zompo MD, Maier W, Hampel H, Pilotto A, Frank-García A, Panza F, Solfrizzi V, Caffarra P, Nacmias B, Perry W, Mayhaus M, Lannfelt L, Hakonarson H, Pichler S, Carrasquillo MM, Ingelsson M, Beekly D, Alvarez V, Zou F, Valladares O, Younkin SG, Coto E, Hamilton-Nelson KL, Gu W, Razquin C, Pastor P, Mateo I, Owen MJ, Faber KM, Jonsson PV, Combarros O, O'Donovan MC, Cantwell LB, Soininen H, Blacker D, Mead S, Mosley TH Jr, Bennett DA, Harris TB, Fratiglioni L, Holmes C, de Bruijn RF, Passmore P, Montine TJ, Bettens K, Rotter JI, Brice A, Morgan K, Foroud TM, Kukull WA, Hannequin D, Powell JF, Nalls MA, Ritchie K, Lunetta KL, Kauwe JS, Boerwinkle E, Riemenschneider M, Boada M, Hiltunen M, Martin ER, Schmidt R, Rujescu D, Dartigues JF, Mayeux R, Tzourio C, Hofman A, Nöthen MM, Graff C, Psaty BM, Haines JL, Lathrop M, Pericak-Vance MA, Launer LJ, Van Broeckhoven C, Farrer LA, van Duijn CM, Ramirez A, Seshadri S, Schellenberg GD, Amouyel P, Williams J; Cardiovascular Health Study (CHS). Gene-wide analysis detects two new susceptibility genes for Alzheimer's disease. PLoS One. 2014 Jun 12;9(6):e94661. PubMed PMID: 24922517; PubMed Central PMCID: PMC4055488.
30. Escott-Price V, Sims R, Bannister C, Harold D, Vronskaya M, Majounie E, Badarinarayan N; GERAD/PERADES; IGAP consortia, Morgan K, Passmore P, Holmes C, Powell J, Brayne C, Gill M, Mead S, Goate A, Cruchaga C, Lambert JC, van Duijn C, Maier W, Ramirez A, Holmans P, Jones L, Hardy J, Seshadri S, Schellenberg GD, Amouyel P, Williams J. Common polygenic variation enhances risk prediction for Alzheimer's disease. Brain. 2015 Dec;138(Pt 12):3673-84. PubMed PMID: 26490334; PubMed Central PMCID: PMC5006219.

31. Fan CC, Banks SJ, Thompson WK, Chen CH, McEvoy LK, Tan CH, Kukull W, Bennett DA, Farrer LA, Mayeux R, Schellenberg GD, Andreassen OA, Desikan R, Dale AM. Sex-dependent autosomal effects on clinical progression of Alzheimer's disease. Brain. 2020 Jul 1;143(7):2272-2280. PMID: 32591829; PMCID: PMC7364740.
32. Fernández MV, Budde J, Del-Aguila JL, Ibañez L, Deming Y, Harari O, Norton J, Morris JC, Goate AM; NIA-LOAD family study group; NCRAD, Cruchaga C. Evaluation of Gene-Based Family-Based Methods to Detect Novel Genes Associated With Familial Late Onset Alzheimer Disease. Front Neurosci. 2018 Apr 4;12:209. doi: 10.3389/fnins.2018.00209. eCollection 2018. PubMed PMID: 29670507; PubMed Central PMCID: PMC5893779.
33. Fernández MV, Kim JH, Budde JP, Black K, Medvedeva A, Saef B, Deming Y, Del-Aguila J, Ibañez L, Dube U, Harari O, Norton J, Chasse R, Morris JC, Goate A; NIA-LOAD family study group; NCRAD, Cruchaga C. Analysis of neurodegenerative Mendelian genes in clinically diagnosed Alzheimer Disease. PLoS Genet. 2017 Nov 1;13(11):e1007045. PubMed PMID: 29091718; PubMed Central PMCID: PMC5683650.
34. Fernández MV, Black K, Carrell D, Saef B, Budde J, Deming Y, Howells B, Del-Aguila JL, Ma S, Bi C, Norton J, Chasse R, Morris J, Goate A, Cruchaga C; NIA-LOAD family study group, NCRAD. SORL1 variants across Alzheimer's disease European American cohorts. Eur J Hum Genet. 2016 Dec;24(12):1828-1830. PubMed PMID: 27650968; PubMed Central PMCID: PMC5117924.
35. Ferrari R, Wang Y, Vandrovcova J, Guelfi S, Witeolar A, Karch CM, Schork AJ, Fan CC, Brewer JB; International FTD-Genomics Consortium (IFGC),; International Parkinson's Disease Genomics Consortium (IPDGC),; International Genomics of Alzheimer's Project (IGAP),, Momeni P, Schellenberg GD, Dillon WP, Sugrue LP, Hess CP, Yokoyama JS, Bonham LW, Rabinovici GD, Miller BL, Andreassen OA, Dale AM, Hardy J, Desikan RS. Genetic architecture of sporadic frontotemporal dementia and overlap with Alzheimer's and Parkinson's diseases. J Neurol Neurosurg Psychiatry. 2017 Feb;88(2):152-164. PubMed PMID: 27899424; PubMed Central PMCID: PMC5237405.
36. Fulton-Howard B, Goate AM, Adelson RP, Koppel J, Gordon ML; Alzheimer's Disease Genetics Consortium, Barzilai N, Atzmon G, Davies P, Freudenberg-Hua Y. Greater effect of polygenic risk score for Alzheimer’s diease among younger cases who are apolipoprotein E-ε4 carriers. Neurobiol Aging. 2021 Mar;99:101.e1-101.e9. PMID: 33164815
37. Ghani M, Reitz C, Cheng R, Vardarajan BN, Jun G, Sato C, Naj A, Rajbhandary R, Wang LS, Valladares O, Lin CF, Larson EB, Graff-Radford NR, Evans D, De Jager PL, Crane PK, Buxbaum JD, Murrell JR, Raj T, Ertekin-Taner N, Logue M, Baldwin CT, Green RC, Barnes LL, Cantwell LB, Fallin MD, Go RC, Griffith PA, Obisesan TO, Manly JJ, Lunetta KL, Kamboh MI, Lopez OL, Bennett DA, Hendrie H, Hall KS, Goate AM, Byrd GS, Kukull WA, Foroud TM, Haines JL, Farrer LA, Pericak-Vance MA, Lee JH, Schellenberg GD, St George-Hyslop P, Mayeux R, Rogaeva E; Alzheimer’s Disease Genetics Consortium. Association of Long Runs of Homozygosity With Alzheimer Disease Among African American Individuals. JAMA Neurol. 2015 Nov;72(11):1313-23. PubMed PMID: 26366463; PubMed Central PMCID: PMC4641052.
38. Gross AL, Sherva R, Mukherjee S, Newhouse S, Kauwe JS, Munsie LM, Waterston LB, Bennett DA, Jones RN, Green RC, Crane PK; Alzheimer's Disease Neuroimaging Initiative; GENAROAD Consortium; AD Genetics Consortium. Calibrating longitudinal cognition in Alzheimer's disease across diverse test batteries and datasets. Neuroepidemiology. 2014;43(3-4):194-205. PubMed PMID: 25402421; PubMed Central PMCID: PMC4297570.
39. Gusareva ES, Twizere JC, Sleegers K, Dourlen P, Abisambra JF, Meier S, Cloyd R, Weiss B, Dermaut B, Bessonov K, van der Lee SJ, Carrasquillo MM, Katsumata Y, Cherkaoui M, Asselbergh B, Ikram MA, Mayeux R, Farrer LA, Haines JL, Pericak-Vance MA, Schellenberg GD; Genetic and Environmental Risk in Alzheimer's Disease 1 consortium (GERAD1); Alzheimer's Disease Genetics Consortium (ADGC); European Alzheimer Disease Initiative Investigators (EADI1 Consortium), Sims R, Williams J, Amouyel P, van Duijn CM, Ertekin-Taner N, Van Broeckhoven C, Dequiedt F, Fardo DW, Lambert JC, Van Steen K. Male-specific epistasis between WWC1 and TLN2 genes is associated with Alzheimer's disease. Neurobiol Aging. 2018 Dec;72:188.e3-188.e12. PubMed PMID: 30201328; PubMed Central PMCID: PMC6769421.
40. Haddick PC, Larson JL, Rathore N, Bhangale TR, Phung QT, Srinivasan K, Hansen DV, Lill JR; Alzheimer’s Disease Genetic Consortium (ADGC), Alzheimer’s Disease Neuroimaging Initiative (ADNI), Pericak-Vance MA, Haines J, Farrer LA, Kauwe JS, Schellenberg GD, Cruchaga C, Goate AM, Behrens TW, Watts RJ, Graham RR, Kaminker JS, van der Brug M. A Common Variant of IL-6R is Associated with Elevated IL-6 Pathway Activity in Alzheimer's Disease Brains. J Alzheimers Dis. 2017;56(3):1037-1054. PubMed PMID: 28106546; PubMed Central PMCID: PMC5667357.
41. Harms M, Benitez BA, Cairns N, Cooper B, Cooper P, Mayo K, Carrell D, Faber K, Williamson J, Bird T, Diaz-Arrastia R, Foroud TM, Boeve BF, Graff-Radford NR, Mayeux R, Chakraverty S, Goate AM, Cruchaga C; NIA-LOAD/NCRAD Family Study Consortium. C9orf72 hexanucleotide repeat expansions in clinical Alzheimer disease. JAMA Neurol. 2013 Jun;70(6):736-41. PubMed PMID: 23588422; PubMed Central PMCID: PMC3681841.

42. Hamshere ML, Holmans PA, Avramopoulos D, Bassett SS, Blacker D, Bertram L, Wiener H, Rochberg N, Tanzi RE, Myers A, Wavrant-De Vrièze F, Go R, Fallin D, Lovestone S, Hardy J, Goate A, O'Donovan M, Williams J, Owen MJ. Genome-wide linkage analysis of 723 affected relative pairs with late-onset Alzheimer's disease. Hum Mol Genet. 2007 Nov 15;16(22):2703-12. PubMed PMID: 17725986.
43. He Z, Zhang D, Renton AE, Li B, Zhao L, Wang GT, Goate AM, Mayeux R, Leal SM. The Rare-Variant Generalized Disequilibrium Test for Association Analysis of Nuclear and Extended Pedigrees with Application to Alzheimer Disease WGS Data. Am J Hum Genet. 2017 Feb 2;100(2):193-204. doi: Erratum in: Am J Hum Genet. 2017 Feb 2;100(2):371. PubMed PMID: 28065470; PubMed Central PMCID: PMC5294711.
44. Herold C, Hooli BV, Mullin K, Liu T, Roehr JT, Mattheisen M, Parrado AR, Bertram L, Lange C, Tanzi RE. Family-based association analyses of imputed genotypes reveal genome-wide significant association of Alzheimer's disease with OSBPL6, PTPRG, and PDCL3. Mol Psychiatry. 2016 Nov;21(11):1608-1612. Epub 2016 Feb 2. PubMed PMID: 26830138; PubMed Central PMCID: PMC4970971.
45. Hohman TJ, Bush WS, Jiang L, Brown-Gentry KD, Torstenson ES, Dudek SM, Mukherjee S, Naj A, Kunkle BW, Ritchie MD, Martin ER, Schellenberg GD, Mayeux R, Farrer LA, Pericak-Vance MA, Haines JL, Thornton-Wells TA; Alzheimer's Disease Genetics Consortium. Discovery of gene-gene interactions across multiple independent data sets of late onset Alzheimer disease from the Alzheimer Disease Genetics Consortium. Neurobiol Aging. 2016 Feb;38:141-150. PubMed PMID: 26827652; PubMed Central PMCID: PMC4735733.
46. Hohman TJ, Cooke-Bailey JN, Reitz C, Jun G, Naj A, Beecham GW, Liu Z, Carney RM, Vance JM, Cuccaro ML, Rajbhandary R, Vardarajan BN, Wang LS, Valladares O, Lin CF, Larson EB, Graff-Radford NR, Evans D, De Jager PL, Crane PK, Buxbaum JD, Murrell JR, Raj T, Ertekin-Taner N, Logue MW, Baldwin CT, Green RC, Barnes LL, Cantwell LB, Fallin MD, Go RC, Griffith P, Obisesan TO, Manly JJ, Lunetta KL, Kamboh MI, Lopez OL, Bennett DA, Hardy J, Hendrie HC, Hall KS, Goate AM, Lang R, Byrd GS, Kukull WA, Foroud TM, Farrer LA, Martin ER, Pericak-Vance MA, Schellenberg GD, Mayeux R, Haines JL, Thornton-Wells TA; Alzheimer Disease Genetics Consortium. Global and local ancestry in African-Americans: Implications for Alzheimer's disease risk. Alzheimers Dement. 2016 Mar;12(3):233-43. PubMed PMID: 26092349; PubMed Central PMCID: PMC4681680.
47. Hohman TJ, Dumitrescu L, Barnes LL, Thambisetty M, Beecham G, Kunkle B, Gifford KA, Bush WS, Chibnik LB, Mukherjee S, De Jager PL, Kukull W, Crane PK, Resnick SM, Keene CD, Montine TJ, Schellenberg GD, Haines JL, Zetterberg H, Blennow K, Larson EB, Johnson SC, Albert M, Bennett DA, Schneider JA, Jefferson AL; Alzheimer’s Disease Genetics Consortium and the Alzheimer’s Disease Neuroimaging Initiative. Sex-Specific Association of Apolipoprotein E With Cerebrospinal Fluid Levels of Tau. JAMA Neurol. 2018 Aug 1;75(8):989-998. PubMed PMID: 29801024; PubMed Central PMCID: PMC6142927.
48. Hollingworth P, Sweet R, Sims R, Harold D, Russo G, Abraham R, Stretton A, Jones N, Gerrish A, Chapman J, Ivanov D, Moskvina V, Lovestone S, Priotsi P, Lupton M, Brayne C, Gill M, Lawlor B, Lynch A, Craig D, McGuinness B, Johnston J, Holmes C, Livingston G, Bass NJ, Gurling H, McQuillin A; GERAD Consortium; National Institute on Aging Late-Onset Alzheimer's Disease Family Study Group, Holmans P, Jones L, Devlin B, Klei L, Barmada MM, Demirci FY, DeKosky ST, Lopez OL, Passmore P, Owen MJ, O'Donovan MC, Mayeux R, Kamboh MI, Williams J. Genome-wide association study of Alzheimer's disease with psychotic symptoms. Mol Psychiatry. 2012 Dec;17(12):1316-27. PubMed PMID: 22005930; PubMed Central PMCID: PMC3272435.
49. Holton P, Ryten M, Nalls M, Trabzuni D, Weale ME, Hernandez D, Crehan H, Gibbs JR, Mayeux R, Haines JL, Farrer LA, Pericak-Vance MA, Schellenberg GD; Alzheimer's Disease Genetics Consortium, Ramirez-Restrepo M, Engel A, Myers AJ, Corneveaux JJ, Huentelman MJ, Dillman A, Cookson MR, Reiman EM, Singleton A, Hardy J, Guerreiro R. Initial assessment of the pathogenic mechanisms of the recently identified Alzheimer risk Loci. Ann Hum Genet. 2013 Mar;77(2):85-105. PubMed PMID: 23360175; PubMed Central PMCID: PMC3578142.
50. International Genomics of Alzheimer's Disease Consortium (IGAP). Convergent genetic and expression data implicate immunity in Alzheimer's disease. Alzheimers Dement. 2015 Jun;11(6):658-71. PubMed PMID: 25533204; PubMed Central PMCID: PMC4672734.
51. Jakobsdottir J, van der Lee SJ, Bis JC, Chouraki V, Li-Kroeger D, Yamamoto S, Grove ML, Naj A, Vronskaya M, Salazar JL, DeStefano AL, Brody JA, Smith AV, Amin N, Sims R, Ibrahim-Verbaas CA, Choi SH, Satizabal CL, Lopez OL, Beiser A, Ikram MA, Garcia ME, Hayward C, Varga TV, Ripatti S, Franks PW, Hallmans G, Rolandsson O, Jansson JH, Porteous DJ, Salomaa V, Eiriksdottir G, Rice KM, Bellen HJ, Levy D, Uitterlinden AG, Emilsson V, Rotter JI, Aspelund T; Cohorts for Heart and Aging Research in Genomic Epidemiology consortium; Alzheimer’s Disease Genetic Consortium; Genetic and Environmental Risk in Alzheimer’s Disease consortium, O'Donnell CJ, Fitzpatrick AL, Launer LJ, Hofman A, Wang LS, Williams J, Schellenberg GD, Boerwinkle E, Psaty BM, Seshadri S, Shulman JM, Gudnason V, van Duijn CM. Rare Functional Variant in TM2D3 is Associated with Late-Onset Alzheimer's Disease. PLoS Genet. 2016 Oct 20;12(10):e1006327. Erratum in: PLoS Genet. 2016 Nov 28;12 (11):e1006456. PubMed PMID: 27764101; PubMed Central PMCID: PMC5072721.
52. Jun G, Asai H, Zeldich E, Drapeau E, Chen C, Chung J, Park JH, Kim S, Haroutunian V, Foroud T, Kuwano R, Haines JL, Pericak-Vance MA, Schellenberg GD, Lunetta KL, Kim JW, Buxbaum JD, Mayeux R, Ikezu T, Abraham CR, Farrer LA. PLXNA4 is associated with Alzheimer disease and modulates tau phosphorylation. Ann Neurol. 2014 Sep;76(3):379-92. PubMed PMID: 25043464; PubMed Central PMCID: PMC4830273.
53. Jun G, Ibrahim-Verbaas CA, Vronskaya M, Lambert JC, Chung J, Naj AC, Kunkle BW, Wang LS, Bis JC, Bellenguez C, Harold D, Lunetta KL, Destefano AL, Grenier-Boley B, Sims R, Beecham GW, Smith AV, Chouraki V, Hamilton-Nelson KL, Ikram MA, Fievet N, Denning N, Martin ER, Schmidt H, Kamatani Y, Dunstan ML, Valladares O, Laza AR, Zelenika D, Ramirez A, Foroud TM, Choi SH, Boland A, Becker T, Kukull WA, van der Lee SJ, Pasquier F, Cruchaga C, Beekly D, Fitzpatrick AL, Hanon O, Gill M, Barber R, Gudnason V, Campion D, Love S, Bennett DA, Amin N, Berr C, Tsolaki M, Buxbaum JD, Lopez OL, Deramecourt V, Fox NC, Cantwell LB, Tárraga L, Dufouil C, Hardy J, Crane PK, Eiriksdottir G, Hannequin D, Clarke R, Evans D, Mosley TH Jr, Letenneur L, Brayne C, Maier W, De Jager P, Emilsson V, Dartigues JF, Hampel H, Kamboh MI, de Bruijn RF, Tzourio C, Pastor P, Larson EB, Rotter JI, O'Donovan MC, Montine TJ, Nalls MA, Mead S, Reiman EM, Jonsson PV, Holmes C, St George-Hyslop PH, Boada M, Passmore P, Wendland JR, Schmidt R, Morgan K, Winslow AR, Powell JF, Carasquillo M, Younkin SG, Jakobsdóttir J, Kauwe JS, Wilhelmsen KC, Rujescu D, Nöthen MM, Hofman A, Jones L; IGAP Consortium, Haines JL, Psaty BM, Van Broeckhoven C, Holmans P, Launer LJ, Mayeux R, Lathrop M, Goate AM, Escott-Price V, Seshadri S, Pericak-Vance MA, Amouyel P, Williams J, van Duijn CM, Schellenberg GD, Farrer LA. A novel Alzheimer disease locus located near the gene encoding tau protein. Mol Psychiatry. 2016 Jan;21(1):108-17. PubMed PMID: 25778476; PubMed Central PMCID: PMC4573764.
54. Jun G, Naj AC, Beecham GW, Wang LS, Buros J, Gallins PJ, Buxbaum JD, Ertekin-Taner N, Fallin MD, Friedland R, Inzelberg R, Kramer P, Rogaeva E, St George-Hyslop P; Alzheimer's Disease Genetics Consortium, Cantwell LB, Dombroski BA, Saykin AJ, Reiman EM, Bennett DA, Morris JC, Lunetta KL, Martin ER, Montine TJ, Goate AM, Blacker D, Tsuang DW, Beekly D, Cupples LA, Hakonarson H, Kukull W, Foroud TM, Haines J, Mayeux R, Farrer LA, Pericak-Vance MA, Schellenberg GD. Meta-analysis confirms CR1, CLU, and PICALM as alzheimer disease risk loci and reveals interactions with APOE genotypes. Arch Neurol. 2010 Dec;67(12):1473-84. Erratum in: Arch Neurol. 2011 Feb;68(2):159. PubMed PMID: 20697030; PubMed Central PMCID: PMC3048805.
55. Jun G, Vardarajan BN, Buros J, Yu CE, Hawk MV, Dombroski BA, Crane PK, Larson EB; Alzheimer's Disease Genetics Consortium, Mayeux R, Haines JL, Lunetta KL, Pericak-Vance MA, Schellenberg GD, Farrer LA. Comprehensive search for Alzheimer disease susceptibility loci in the APOE region. Arch Neurol. 2012 Oct;69(10):1270-9. PubMed PMID: 22869155; PubMed Central PMCID: PMC3579659.
56. Jun GR, Chung J, Mez J, Barber R, Beecham GW, Bennett DA, Buxbaum JD, Byrd GS, Carrasquillo MM, Crane PK, Cruchaga C, De Jager P, Ertekin-Taner N, Evans D, Fallin MD, Foroud TM, Friedland RP, Goate AM, Graff-Radford NR, Hendrie H, Hall KS, Hamilton-Nelson KL, Inzelberg R, Kamboh MI, Kauwe JSK, Kukull WA, Kunkle BW, Kuwano R, Larson EB, Logue MW, Manly JJ, Martin ER, Montine TJ, Mukherjee S, Naj A, Reiman EM, Reitz C, Sherva R, St George-Hyslop PH, Thornton T, Younkin SG, Vardarajan BN, Wang LS, Wendlund JR, Winslow AR; Alzheimer's Disease Genetics Consortium, Haines J, Mayeux R, Pericak-Vance MA, Schellenberg G, Lunetta KL, Farrer LA. Transethnic genome-wide scan identifies novel Alzheimer's disease loci. Alzheimers Dement. 2017 Jul;13(7):727-738. PubMed PMID: 28183528; PubMed Central PMCID: PMC5496797.
57. Karch CM, Ezerskiy LA, Bertelsen S; Alzheimer’s Disease Genetics Consortium (ADGC), Goate AM. Alzheimer's Disease Risk Polymorphisms Regulate Gene Expression in the ZCWPW1 and the CELF1 Loci. PLoS One. 2016 Feb 26;11(2):e0148717. PubMed PMID: 26919393; PubMed Central PMCID: PMC4769299.
58. Kessler MD, Loesch DP, Perry JA, Heard-Costa NL, Taliun D, Cade BE, Wang H, Daya M, Ziniti J, Datta S, Celedón JC, Soto-Quiros ME, Avila L, Weiss ST, Barnes K, Redline SS, Vasan RS, Johnson AD, Mathias RA, Hernandez R, Wilson JG, Nickerson DA, Abecasis G, Browning SR, Zöllner S, O'Connell JR, Mitchell BD; National Heart, Lung, and Blood Institute Trans-Omics for Precision Medicine (TOPMed) Consortium; TOPMed Population Genetics Working Group, O'Connor TD. De novo mutations across 1,465 diverse genomes reveal mutational insights and reductions in the Amish founder population. Proc Natl Acad Sci U S A. 2020 Feb 4;117(5):2560-2569 PMID: 31964835; PMCID: PMC7007577.
59. Kohli MA, Cukier HN, Hamilton-Nelson KL, Rolati S, Kunkle BW, Whitehead PL, Züchner SL, Farrer LA, Martin ER, Beecham GW, Haines JL, Vance JM, Cuccaro ML, Gilbert JR, Schellenberg GD, Carney RM, Pericak-Vance MA. Segregation of a rare TTC3 variant in an extended family with late-onset Alzheimer disease. Neurol Genet. 2016 Jan 14;2(1):e41. PubMed PMID: 27066578; PubMed Central PMCID: PMC4817909.
60. Kirola L, Budde JP, Wang F, Norton J, Morris JC, NIA-LOAD Family Study Group, NCRAD, The ADSP project, Cruchaga C, Fernandez MV. Lack of evidence supporting a role for DPP6 sequence variants in Alzheimer’s disease in the European American population. Acta Neuropathol 2021; 141, 623–624. PMID:33591372
61. Kunkle BW, Grenier-Boley B, Sims R, Bis JC, Damotte V, Naj AC, Boland A, Vronskaya M, van der Lee SJ, Amlie-Wolf A, Bellenguez C, Frizatti A, Chouraki V, Martin ER, Sleegers K, Badarinarayan N, Jakobsdottir J, Hamilton-Nelson KL, Moreno-Grau S, Olaso R, Raybould R, Chen Y, Kuzma AB, Hiltunen M, Morgan T, Ahmad S, Vardarajan BN, Epelbaum J, Hoffmann P, Boada M, Beecham GW, Garnier JG, Harold D, Fitzpatrick AL, Valladares O, Moutet ML, Gerrish A, Smith AV, Qu L, Bacq D, Denning N, Jian X, Zhao Y, Del Zompo M, Fox NC, Choi SH, Mateo I, Hughes JT, Adams HH, Malamon J, Sanchez-Garcia F, Patel Y, Brody JA, Dombroski BA, Naranjo MCD, Daniilidou M, Eiriksdottir G, Mukherjee S, Wallon D, Uphill J, Aspelund T, Cantwell LB, Garzia F, Galimberti D, Hofer E, Butkiewicz M, Fin B, Scarpini E, Sarnowski C, Bush WS, Meslage S, Kornhuber J, White CC, Song Y, Barber RC, Engelborghs S, Sordon S, Voijnovic D, Adams PM, Vandenberghe R, Mayhaus M, Cupples LA, Albert MS, De Deyn PP, Gu W, Himali JJ, Beekly D, Squassina A, Hartmann AM, Orellana A, Blacker D, Rodriguez-Rodriguez E, Lovestone S, Garcia ME, Doody RS, Munoz-Fernadez C, Sussams R, Lin H, Fairchild TJ, Benito YA, Holmes C, Karamujić-Čomić H, Frosch MP, Thonberg H, Maier W, Roshchupkin G, Ghetti B, Giedraitis V, Kawalia A, Li S, Huebinger RM, Kilander L, Moebus S, Hernández I, Kamboh MI, Brundin R, Turton J, Yang Q, Katz MJ, Concari L, Lord J, Beiser AS, Keene CD, Helisalmi S, Kloszewska I, Kukull WA, Koivisto AM, Lynch A, Tarraga L, Larson EB, Haapasalo A, Lawlor B, Mosley TH, Lipton RB, Solfrizzi V, Gill M, Longstreth WT Jr, Montine TJ, Frisardi V, Diez-Fairen M, Rivadeneira F, Petersen RC, Deramecourt V, Alvarez I, Salani F, Ciaramella A, Boerwinkle E, Reiman EM, Fievet N, Rotter JI, Reisch JS, Hanon O, Cupidi C, Andre Uitterlinden AG, Royall DR, Dufouil C, Maletta RG, de Rojas I, Sano M, Brice A, Cecchetti R, George-Hyslop PS, Ritchie K, Tsolaki M, Tsuang DW, Dubois B, Craig D, Wu CK, Soininen H, Avramidou D, Albin RL, Fratiglioni L, Germanou A, Apostolova LG, Keller L, Koutroumani M, Arnold SE, Panza F, Gkatzima O, Asthana S, Hannequin D, Whitehead P, Atwood CS, Caffarra P, Hampel H, Quintela I, Carracedo Á, Lannfelt L, Rubinsztein DC, Barnes LL, Pasquier F, Frölich L, Barral S, McGuinness B, Beach TG, Johnston JA, Becker JT, Passmore P, Bigio EH, Schott JM, Bird TD, Warren JD, Boeve BF, Lupton MK, Bowen JD, Proitsi P, Boxer A, Powell JF, Burke JR, Kauwe JSK, Burns JM, Mancuso M, Buxbaum JD, Bonuccelli U, Cairns NJ, McQuillin A, Cao C, Livingston G, Carlson CS, Bass NJ, Carlsson CM, Hardy J, Carney RM, Bras J, Carrasquillo MM, Guerreiro R, Allen M, Chui HC, Fisher E, Masullo C, Crocco EA, DeCarli C, Bisceglio G, Dick M, Ma L, Duara R, Graff-Radford NR, Evans DA, Hodges A, Faber KM, Scherer M, Fallon KB, Riemenschneider M, Fardo DW, Heun R, Farlow MR, Kölsch H, Ferris S, Leber M, Foroud TM, Heuser I, Galasko DR, Giegling I, Gearing M, Hüll M, Geschwind DH, Gilbert JR, Morris J, Green RC, Mayo K, Growdon JH, Feulner T, Hamilton RL, Harrell LE, Drichel D, Honig LS, Cushion TD, Huentelman MJ, Hollingworth P, Hulette CM, Hyman BT, Marshall R, Jarvik GP, Meggy A, Abner E, Menzies GE, Jin LW, Leonenko G, Real LM, Jun GR, Baldwin CT, Grozeva D, Karydas A, Russo G, Kaye JA, Kim R, Jessen F, Kowall NW, Vellas B, Kramer JH, Vardy E, LaFerla FM, Jöckel KH, Lah JJ, Dichgans M, Leverenz JB, Mann D, Levey AI, Pickering-Brown S, Lieberman AP, Klopp N, Lunetta KL, Wichmann HE, Lyketsos CG, Morgan K, Marson DC, Brown K, Martiniuk F, Medway C, Mash DC, Nöthen MM, Masliah E, Hooper NM, McCormick WC, Daniele A, McCurry SM, Bayer A, McDavid AN, Gallacher J, McKee AC, van den Bussche H, Mesulam M, Brayne C, Miller BL, Riedel-Heller S, Miller CA, Miller JW, Al-Chalabi A, Morris JC, Shaw CE, Myers AJ, Wiltfang J, O'Bryant S, Olichney JM, Alvarez V, Parisi JE, Singleton AB, Paulson HL, Collinge J, Perry WR, Mead S, Peskind E, Cribbs DH, Rossor M, Pierce A, Ryan NS, Poon WW, Nacmias B, Potter H, Sorbi S, Quinn JF, Sacchinelli E, Raj A, Spalletta G, Raskind M, Caltagirone C, Bossù P, Orfei MD, Reisberg B, Clarke R, Reitz C, Smith AD, Ringman JM, Warden D, Roberson ED, Wilcock G, Rogaeva E, Bruni AC, Rosen HJ, Gallo M, Rosenberg RN, Ben-Shlomo Y, Sager MA, Mecocci P, Saykin AJ, Pastor P, Cuccaro ML, Vance JM, Schneider JA, Schneider LS, Slifer S, Seeley WW, Smith AG, Sonnen JA, Spina S, Stern RA, Swerdlow RH, Tang M, Tanzi RE, Trojanowski JQ, Troncoso JC, Van Deerlin VM, Van Eldik LJ, Vinters HV, Vonsattel JP, Weintraub S, Welsh-Bohmer KA, Wilhelmsen KC, Williamson J, Wingo TS, Woltjer RL, Wright CB, Yu CE, Yu L, Saba Y, Pilotto A, Bullido MJ, Peters O, Crane PK, Bennett D, Bosco P, Coto E, Boccardi V, De Jager PL, Lleo A, Warner N, Lopez OL, Ingelsson M, Deloukas P, Cruchaga C, Graff C, Gwilliam R, Fornage M, Goate AM, Sanchez-Juan P, Kehoe PG, Amin N, Ertekin-Taner N, Berr C, Debette S, Love S, Launer LJ, Younkin SG, Dartigues JF, Corcoran C, Ikram MA, Dickson DW, Nicolas G, Campion D, Tschanz J, Schmidt H, Hakonarson H, Clarimon J, Munger R, Schmidt R, Farrer LA, Van Broeckhoven C, C O'Donovan M, DeStefano AL, Jones L, Haines JL, Deleuze JF, Owen MJ, Gudnason V, Mayeux R, Escott-Price V, Psaty BM, Ramirez A, Wang LS, Ruiz A, van Duijn CM, Holmans PA, Seshadri S, Williams J, Amouyel P, Schellenberg GD, Lambert JC, Pericak-Vance MA; Alzheimer Disease Genetics Consortium (ADGC),; European Alzheimer’s Disease Initiative (EADI),; Cohorts for Heart and Aging Research in Genomic Epidemiology Consortium (CHARGE),; Genetic and Environmental Risk in AD/Defining Genetic, Polygenic and Environmental Risk for Alzheimer’s Disease Consortium (GERAD/PERADES),. Genetic meta-analysis of diagnosed Alzheimer's disease identifies new risk loci and implicates Aβ, tau, immunity and lipid processing. Nat Genet. 2019 Mar;51(3):414-430. Erratum in: Nat Genet. 2019 Sep;51(9):1423-1424. PubMed PMID: 30820047; PubMed Central PMCID: PMC6463297.
62. Kunkle BW, Jaworski J, Barral S, Vardarajan B, Beecham GW, Martin ER, Cantwell LS, Partch A, Bird TD, Raskind WH, DeStefano AL, Carney RM, Cuccaro M, Vance JM, Farrer LA, Goate AM, Foroud T, Mayeux RP, Schellenberg GD, Haines JL, Pericak-Vance MA. Genome-wide linkage analyses of non-Hispanic white families identify novel loci for familial late-onset Alzheimer's disease. Alzheimers Dement. 2016 Jan;12(1):2-10. PubMed PMID: 26365416; PubMed Central PMCID: PMC4717829.
63. Kunkle BW, Vardarajan BN, Naj AC, Whitehead PL, Rolati S, Slifer S, Carney RM, Cuccaro ML, Vance JM, Gilbert JR, Wang LS, Farrer LA, Reitz C, Haines JL, Beecham GW, Martin ER, Schellenberg GD, Mayeux RP, Pericak-Vance MA. Early-Onset Alzheimer Disease and Candidate Risk Genes Involved in Endolysosomal Transport. JAMA Neurol. 2017 Sep 1;74(9):1113-1122. PubMed PMID: 28738127; PubMed Central PMCID: PMC5691589.

64. Kunkle BW, Schmidt M, Klein HU, Naj AC, Hamilton-Nelson KL, Larson EB, Evans DA, De Jager PL, Crane PK, Buxbaum JD, Ertekin-Taner N, Barnes LL, Fallin MD, Manly JJ, Go RCP, Obisesan TO, Kamboh MI, Bennett DA, Hall KS, Goate AM, Foroud TM, Martin ER, Wang LS, Byrd GS, Farrer LA, Haines JL, Schellenberg GD, Mayeux R, Pericak-Vance MA, Reitz C; Alzheimer’s Disease Genetics Consortium (ADGC), Graff-Radford NR, Martinez I, Ayodele T, Logue MW, Cantwell LB, Jean-Francois M, Kuzma AB, Adams LD, Vance JM, Cuccaro ML, Chung J, Mez J, Lunetta KL, Jun GR, Lopez OL, Hendrie HC, Reiman EM, Kowall NW, Leverenz JB, Small SA, Levey AI, Golde TE, Saykin AJ, Starks TD, Albert MS, Hyman BT, Petersen RC, Sano M, Wisniewski T, Vassar R, Kaye JA, Henderson VW, DeCarli C, LaFerla FM, Brewer JB, Miller BL, Swerdlow RH, Van Eldik LJ, Paulson HL, Trojanowski JQ, Chui HC, Rosenberg RN, Craft S, Grabowski TJ, Asthana S, Morris JC, Strittmatter SM, Kukull WA. Novel Alzheimer Disease Risk Loci and Pathways in African American Individuals Using the African Genome Resources Panel: A Meta-analysis. JAMA Neurol. 2020 Oct 19:e203536. PMID: 33074286; PMCID: PMC7573798.
65. Lambert JC, Ibrahim-Verbaas CA, Harold D, Naj AC, Sims R, Bellenguez C, DeStafano AL, Bis JC, Beecham GW, Grenier-Boley B, Russo G, Thorton-Wells TA, Jones N, Smith AV, Chouraki V, Thomas C, Ikram MA, Zelenika D, Vardarajan BN, Kamatani Y, Lin CF, Gerrish A, Schmidt H, Kunkle B, Dunstan ML, Ruiz A, Bihoreau MT, Choi SH, Reitz C, Pasquier F, Cruchaga C, Craig D, Amin N, Berr C, Lopez OL, De Jager PL, Deramecourt V, Johnston JA, Evans D, Lovestone S, Letenneur L, Morón FJ, Rubinsztein DC, Eiriksdottir G, Sleegers K, Goate AM, Fiévet N, Huentelman MW, Gill M, Brown K, Kamboh MI, Keller L, Barberger-Gateau P, McGuiness B, Larson EB, Green R, Myers AJ, Dufouil C, Todd S, Wallon D, Love S, Rogaeva E, Gallacher J, St George-Hyslop P, Clarimon J, Lleo A, Bayer A, Tsuang DW, Yu L, Tsolaki M, Bossù P, Spalletta G, Proitsi P, Collinge J, Sorbi S, Sanchez-Garcia F, Fox NC, Hardy J, Deniz Naranjo MC, Bosco P, Clarke R, Brayne C, Galimberti D, Mancuso M, Matthews F; European Alzheimer's Disease Initiative (EADI); Genetic and Environmental Risk in Alzheimer's Disease; Alzheimer's Disease Genetic Consortium; Cohorts for Heart and Aging Research in Genomic Epidemiology, Moebus S, Mecocci P, Del Zompo M, Maier W, Hampel H, Pilotto A, Bullido M, Panza F, Caffarra P, Nacmias B, Gilbert JR, Mayhaus M, Lannefelt L, Hakonarson H, Pichler S, Carrasquillo MM, Ingelsson M, Beekly D, Alvarez V, Zou F, Valladares O, Younkin SG, Coto E, Hamilton-Nelson KL, Gu W, Razquin C, Pastor P, Mateo I, Owen MJ, Faber KM, Jonsson PV, Combarros O, O'Donovan MC, Cantwell LB, Soininen H, Blacker D, Mead S, Mosley TH Jr, Bennett DA, Harris TB, Fratiglioni L, Holmes C, de Bruijn RF, Passmore P, Montine TJ, Bettens K, Rotter JI, Brice A, Morgan K, Foroud TM, Kukull WA, Hannequin D, Powell JF, Nalls MA, Ritchie K, Lunetta KL, Kauwe JS, Boerwinkle E, Riemenschneider M, Boada M, Hiltuenen M, Martin ER, Schmidt R, Rujescu D, Wang LS, Dartigues JF, Mayeux R, Tzourio C, Hofman A, Nöthen MM, Graff C, Psaty BM, Jones L, Haines JL, Holmans PA, Lathrop M, Pericak-Vance MA, Launer LJ, Farrer LA, van Duijn CM, Van Broeckhoven C, Moskvina V, Seshadri S, Williams J, Schellenberg GD, Amouyel P. Meta-analysis of 74,046 individuals identifies 11 new susceptibility loci for Alzheimer's disease. Nat Genet. 2013 Dec;45(12):1452-8. PubMed PMID: 24162737; PubMed Central PMCID: PMC3896259.
66. Lancour D, Naj A, Mayeux R, Haines JL, Pericak-Vance MA, Schellenberg GD, Crovella M, Farrer LA, Kasif S. One for all and all for One: Improving replication of genetic studies through network diffusion. PLoS Genet. 2018 Apr 23;14(4):e1007306. PubMed PMID: 29684019; PubMed Central PMCID: PMC5933817.
67. Lancour D, Dupuis J, Mayeux R, Haines JL, Pericak-Vance MA, Schellenberg GC, Crovella M, Farrer LA, Kasif S. Analysis of brain region-specific co-expression networks reveals clustering of established and novel genes associated with Alzheimer disease. Alzheimers Res Ther. 2020 Sep 2;12(1):103. PMID: 32878640; PMCID: PMC7469336.
68. Lee JH, Cheng R, Graff-Radford N, Foroud T, Mayeux R; National Institute on Aging Late-Onset Alzheimer's Disease Family Study Group. Analyses of the National Institute on Aging Late-Onset Alzheimer's Disease Family Study: implication of additional loci. Arch Neurol. 2008 Nov;65(11):1518-26. PubMed PMID: 19001172; PubMed Central PMCID: PMC2694670.
69. Lee JH, Cheng R, Vardarajan B, Lantigua R, Reyes-Dumeyer D, Ortmann W, Graham RR, Bhangale T, Behrens TW, Medrano M, Jiménez-Velázquez IZ, Mayeux R. Genetic Modifiers of Age at Onset in Carriers of the G206A Mutation in PSEN1 With Familial Alzheimer Disease Among Caribbean Hispanics. JAMA Neurol. 2015 Sep;72(9):1043-51. PubMed PMID: 26214276; PubMed Central PMCID: PMC5010776
70. Lee S, Zhou X, Gao Y, Vardarajan B, Reyes-Dumeyer D, Rajan KB, Wilson RS, Evans DA, Besser LM, Kukull WA, Bennett DA, Brickman AM, Schupf N, Mayeux R, Barral S. Episodic memory performance in a multi-ethnic longitudinal study of 13,037 elderly. PLoS One. 2018 Nov 21;13(11):e0206803. PubMed PMID: 30462667; PubMed Central PMCID: PMC6248922.
71. Logue MW, Lancour D, Farrell J, Simkina I, Fallin MD, Lunetta KL, Farrer LA. Targeted sequencing of Alzheimer’s disease genes in African Americans implicates novel risk variants. Front Neurosci. 2018 Aug 27;12:592. PMID: 30210277.
72. Logue MW, Schu M, Vardarajan BN, Farrell J, Bennett DA, Buxbaum JD, Byrd GS, Ertekin-Taner N, Evans D, Foroud T, Goate A, Graff-Radford NR, Kamboh MI, Kukull WA, Manly JJ; Alzheimer Disease Genetics Consortium; Alzheimer Disease Genetics Consortium. Two rare AKAP9 variants are associated with Alzheimer's disease in African Americans. Alzheimers Dement. 2014 Nov;10(6):609-618.e11. PubMed PMID: 25172201; PubMed Central PMCID: PMC4253055.
73. Ma Y, Jun GR, Zhang X, Chung J, Naj AC, Chen Y, Bellenguez C, Hamilton-Nelson K, Martin ER, Kunkle BW, Bis JC, Debette S, DeStefano AL, Fornage M, Nicolas G, van Duijn C, Bennett DA, De Jager PL, Mayeux R, Haines JL, Pericak-Vance MA, Seshadri S, Lambert JC, Schellenberg GD, Lunetta KL, Farrer LA; Alzheimer’s Disease Sequencing Project and Alzheimer’s Disease Exome Sequencing–France Project. Analysis of Whole-Exome Sequencing Data for Alzheimer Disease Stratified by APOE Genotype. JAMA Neurol. 2019 Jun 10. PubMed PMID: 31180460; PubMed Central PMCID: PMC6563544.
74. Ma Y, Jun GR, Chung J, Zhang X, Kunkle BW, Naj AC, White CC, Bennett DA, De Jager PL; Alzheimer’s Disease Genetics Consortium, Mayeux R, Haines JL, Pericak- Vance MA, Schellenberg GD, Farrer LA, Lunetta KL. CpG-related SNPs in the MS4A region have a dose-dependent effect on risk of late-onset Alzheimer disease. Aging Cell. 2019 Aug;18(4):e12964.. PMID: 31144443; PMCID: PMC6612647.
75. Mez J, Chung J, Jun G, Kriegel J, Bourlas AP, Sherva R, Logue MW, Barnes LL, Bennett DA, Buxbaum JD, Byrd GS, Crane PK, Ertekin-Taner N, Evans D, Fallin MD, Foroud T, Goate A, Graff-Radford NR, Hall KS, Kamboh MI, Kukull WA, Larson EB, Manly JJ; Alzheimer's Disease Genetics Consortium, Haines JL, Mayeux R, Pericak-Vance MA, Schellenberg GD, Lunetta KL, Farrer LA. Two novel loci, COBL and SLC10A2, for Alzheimer's disease in African Americans. Alzheimers Dement. 2017 Feb;13(2):119-129. PubMed PMID: 27770636; PubMed Central PMCID: PMC5318231.
76. Miyashita A, Koike A, Jun G, Wang LS, Takahashi S, Matsubara E, Kawarabayashi T, Shoji M, Tomita N, Arai H, Asada T, Harigaya Y, Ikeda M, Amari M, Hanyu H, Higuchi S, Ikeuchi T, Nishizawa M, Suga M, Kawase Y, Akatsu H, Kosaka K, Yamamoto T, Imagawa M, Hamaguchi T, Yamada M, Morihara T, Takeda M, Takao T, Nakata K, Fujisawa Y, Sasaki K, Watanabe K, Nakashima K, Urakami K, Ooya T, Takahashi M, Yuzuriha T, Serikawa K, Yoshimoto S, Nakagawa R, Kim JW, Ki CS, Won HH, Na DL, Seo SW, Mook-Jung I; Alzheimer Disease Genetics Consortium, St George-Hyslop P, Mayeux R, Haines JL, Pericak-Vance MA, Yoshida M, Nishida N, Tokunaga K, Yamamoto K, Tsuji S, Kanazawa I, Ihara Y, Schellenberg GD, Farrer LA, Kuwano R. SORL1 is genetically associated with late-onset Alzheimer's disease in Japanese, Koreans and Caucasians. PLoS One. 2013;8(4):e58618. Erratum in: PLoS One. 2013;8(7). doi:10.1371/annotation/fcb56ea7-d32a-4e45-818d-39cef330c731. Moriaha, Takashi [corrected to Morihara, Takashi]. PubMed PMID: 23565137; PubMed Central PMCID: PMC3614978.
77. Moreno-Grau S, Fernández MV, de Rojas I, Garcia-González P, Hernández I, Farias F, Budde JP, Quintela I, Madrid L, González-Pérez A, Montrreal L, Alarcón-Martín E, Alegret M, Maroñas O, Pineda JA, Macías J; GR@ACE study group; DEGESCO consortium, Marquié M, Valero S, Benaque A, Clarimón J, Bullido MJ, García-Ribas G, Pástor P, Sánchez-Juan P, Álvarez V, Piñol-Ripoll G, García-Alberca JM, Royo JL, Franco-Macías E, Mir P, Calero M, Medina M, Rábano A, Ávila J, Antúnez C, Real LM, Orellana A, Carracedo Á, Sáez ME, Tárraga L, Boada M, Cruchaga C, Ruiz A; Alzheimer’s Disease Neuroimaging Initiative. Long runs of homozygosity are associated with Alzheimer’s disease. Transl Psychiatry 2021; 11: 142. PMID: 33627629; PMCID: PMC7904832
78. Nafikov RA, Nato AQ Jr, Sohi H, Wang B, Brown L, Horimoto AR, Vardarajan BN, Barral SM, Tosto G, Mayeux RP, Thornton TA, Blue E, Wijsman EM. Analysis of pedigree data in populations with multiple ancestries: Strategies for dealing with admixture in Caribbean Hispanic families from the ADSP. Genet Epidemiol. 2018 Sep;42(6):500-515. PubMed PMID: 29862559; PubMed Central PMCID: PMC6160322.
79. Naj AC, Jun G, Beecham GW, Wang LS, Vardarajan BN, Buros J, Gallins PJ, Buxbaum JD, Jarvik GP, Crane PK, Larson EB, Bird TD, Boeve BF, Graff-Radford NR, De Jager PL, Evans D, Schneider JA, Carrasquillo MM, Ertekin-Taner N, Younkin SG, Cruchaga C, Kauwe JS, Nowotny P, Kramer P, Hardy J, Huentelman MJ, Myers AJ, Barmada MM, Demirci FY, Baldwin CT, Green RC, Rogaeva E, St George-Hyslop P, Arnold SE, Barber R, Beach T, Bigio EH, Bowen JD, Boxer A, Burke JR, Cairns NJ, Carlson CS, Carney RM, Carroll SL, Chui HC, Clark DG, Corneveaux J, Cotman CW, Cummings JL, DeCarli C, DeKosky ST, Diaz-Arrastia R, Dick M, Dickson DW, Ellis WG, Faber KM, Fallon KB, Farlow MR, Ferris S, Frosch MP, Galasko DR, Ganguli M, Gearing M, Geschwind DH, Ghetti B, Gilbert JR, Gilman S, Giordani B, Glass JD, Growdon JH, Hamilton RL, Harrell LE, Head E, Honig LS, Hulette CM, Hyman BT, Jicha GA, Jin LW, Johnson N, Karlawish J, Karydas A, Kaye JA, Kim R, Koo EH, Kowall NW, Lah JJ, Levey AI, Lieberman AP, Lopez OL, Mack WJ, Marson DC, Martiniuk F, Mash DC, Masliah E, McCormick WC, McCurry SM, McDavid AN, McKee AC, Mesulam M, Miller BL, Miller CA, Miller JW, Parisi JE, Perl DP, Peskind E, Petersen RC, Poon WW, Quinn JF, Rajbhandary RA, Raskind M, Reisberg B, Ringman JM, Roberson ED, Rosenberg RN, Sano M, Schneider LS, Seeley W, Shelanski ML, Slifer MA, Smith CD, Sonnen JA, Spina S, Stern RA, Tanzi RE, Trojanowski JQ, Troncoso JC, Van Deerlin VM, Vinters HV, Vonsattel JP, Weintraub S, Welsh-Bohmer KA, Williamson J, Woltjer RL, Cantwell LB, Dombroski BA, Beekly D, Lunetta KL, Martin ER, Kamboh MI, Saykin AJ, Reiman EM, Bennett DA, Morris JC, Montine TJ, Goate AM, Blacker D, Tsuang DW, Hakonarson H, Kukull WA, Foroud TM, Haines JL, Mayeux R, Pericak-Vance MA, Farrer LA, Schellenberg GD. Common variants at MS4A4/MS4A6E, CD2AP, CD33 and EPHA1 are associated with late-onset Alzheimer's disease. Nat Genet. 2011 May;43(5):436-41. PubMed PMID: 21460841; PubMed Central PMCID: PMC3090745.
80. Naj AC, Jun G, Reitz C, Kunkle BW, Perry W, Park YS, Beecham GW, Rajbhandary RA, Hamilton-Nelson KL, Wang LS, Kauwe JS, Huentelman MJ, Myers AJ, Bird TD, Boeve BF, Baldwin CT, Jarvik GP, Crane PK, Rogaeva E, Barmada MM, Demirci FY, Cruchaga C, Kramer PL, Ertekin-Taner N, Hardy J, Graff-Radford NR, Green RC, Larson EB, St George-Hyslop PH, Buxbaum JD, Evans DA, Schneider JA, Lunetta KL, Kamboh MI, Saykin AJ, Reiman EM, De Jager PL, Bennett DA, Morris JC, Montine TJ, Goate AM, Blacker D, Tsuang DW, Hakonarson H, Kukull WA, Foroud TM, Martin ER, Haines JL, Mayeux RP, Farrer LA, Schellenberg GD, Pericak-Vance MA; Alzheimer Disease Genetics Consortium, Albert MS, Albin RL, Apostolova LG, Arnold SE, Barber R, Barnes LL, Beach TG, Becker JT, Beekly D, Bigio EH, Bowen JD, Boxer A, Burke JR, Cairns NJ, Cantwell LB, Cao C, Carlson CS, Carney RM, Carrasquillo MM, Carroll SL, Chui HC, Clark DG, Corneveaux J, Cribbs DH, Crocco EA, DeCarli C, DeKosky ST, Dick M, Dickson DW, Duara R, Faber KM, Fallon KB, Farlow MR, Ferris S, Frosch MP, Galasko DR, Ganguli M, Gearing M, Geschwind DH, Ghetti B, Gilbert JR, Glass JD, Growdon JH, Hamilton RL, Harrell LE, Head E, Honig LS, Hulette CM, Hyman BT, Jicha GA, Jin LW, Karydas A, Kaye JA, Kim R, Koo EH, Kowall NW, Kramer JH, LaFerla FM, Lah JJ, Leverenz JB, Levey AI, Li G, Lieberman AP, Lin CF, Lopez OL, Lyketsos CG, Mack WJ, Martiniuk F, Mash DC, Masliah E, McCormick WC, McCurry SM, McDavid AN, McKee AC, Mesulam M, Miller BL, Miller CA, Miller JW, Murrell JR, Olichney JM, Pankratz VS, Parisi JE, Paulson HL, Peskind E, Petersen RC, Pierce A, Poon WW, Potter H, Quinn JF, Raj A, Raskind M, Reisberg B, Ringman JM, Roberson ED, Rosen HJ, Rosenberg RN, Sano M, Schneider LS, Seeley WW, Smith AG, Sonnen JA, Spina S, Stern RA, Tanzi RE, Thornton-Wells TA, Trojanowski JQ, Troncoso JC, Valladares O, Van Deerlin VM, Van Eldik LJ, Vardarajan BN, Vinters HV, Vonsattel JP, Weintraub S, Welsh-Bohmer KA, Williamson J, Wishnek S, Woltjer RL, Wright CB, Younkin SG, Yu CE, Yu L. Effects of multiple genetic loci on age at onset in late-onset Alzheimer disease: a genome-wide association study. JAMA Neurol. 2014 Nov;71(11):1394-404. JAMA Neurol. 2014 Nov;71(11):1457. PubMed PMID: 25199842; PubMed Central PMCID: PMC4314944.
81. Naj AC, Schellenberg GD; Alzheimer's Disease Genetics Consortium (ADGC). Genomic variants, genes, and pathways of Alzheimer's disease: An overview. Am J Med Genet B Neuropsychiatr Genet. 2017 Jan;174(1):5-26. PubMed PMID: 27943641; PubMed Central PMCID: PMC6179157.
82. Naj AC, Lin H, Vardarajan BN, White S, Lancour D, Ma Y, Schmidt M, Sun F, Butkiewicz M, Bush WS, Kunkle BW, Malamon J, Amin N, Choi SH, Hamilton-Nelson KL, van der Lee SJ, Gupta N, Koboldt DC, Saad M, Wang B, Nato AQ Jr, Sohi HK, Kuzma A; Alzheimer's Disease Sequencing Project (ADSP), Wang LS, Cupples LA, van Duijn C, Seshadri S, Schellenberg GD, Boerwinkle E, Bis JC, Dupuis J, Salerno WJ, Wijsman EM, Martin ER, DeStefano AL. Quality control and integration of genotypes from two calling pipelines for whole genome sequence data in the Alzheimer's disease sequencing project. Genomics. 2019 Jul;111(4):808-818. PMID: 29857119
83. Nelson PT, Estus S, Abner EL, Parikh I, Malik M, Neltner JH, Ighodaro E, Wang WX, Wilfred BR, Wang LS, Kukull WA, Nandakumar K, Farman ML, Poon WW, Corrada MM, Kawas CH, Cribbs DH, Bennett DA, Schneider JA, Larson EB, Crane PK, Valladares O, Schmitt FA, Kryscio RJ, Jicha GA, Smith CD, Scheff SW, Sonnen JA, Haines JL, Pericak-Vance MA, Mayeux R, Farrer LA, Van Eldik LJ, Horbinski C, Green RC, Gearing M, Poon LW, Kramer PL, Woltjer RL, Montine TJ, Partch AB, Rajic AJ, Richmire K, Monsell SE; Alzheimer’ Disease Genetic Consortium, Schellenberg GD, Fardo DW. ABCC9 gene polymorphism is associated with hippocampal sclerosis of aging pathology. Acta Neuropathol. 2014;127(6):825-43. PubMed PMID: 24770881; PubMed Central PMCID: PMC4113197.
84. Olive C, Ibanez L, Farias FHG, Wang F, Budde JP, Norton JB, Gentsch J, Morris JC, Li Z, Dube U, Del-Aguila J, Bergmann K, Bradley J, Benitez BA, Harari O, Fagan A, Ances B, Cruchaga C, Fernandez MV. Examination of the Effect of Rare Variants in TREM2, ABI3, and PLCG2 in LOAD Through Multiple Phenotypes. J Alzheimers Dis. 2020; 77: 1469-1482. PMID:32894242; PMCID: PMC7927150.
85. Østergaard SD, Mukherjee S, Sharp SJ, Proitsi P, Lotta LA, Day F, Perry JR, Boehme KL, Walter S, Kauwe JS, Gibbons LE; Alzheimer’s Disease Genetics Consortium; GERAD1 Consortium; EPIC-InterAct Consortium, Larson EB, Powell JF, Langenberg C, Crane PK, Wareham NJ, Scott RA. Associations between Potentially Modifiable Risk Factors and Alzheimer Disease: A Mendelian Randomization Study. PLoS Med. 2015 Jun 16;12(6):e1001841; discussion e1001841. eCollection 2015 Jun. PubMed PMID: 26079503; PubMed Central PMCID: PMC4469461.
86. Peloso GM, van der Lee SJ; International Genomics of Alzheimer's Project (IGAP), Destefano AL, Seshardi S. Genetically elevated high-density lipoprotein cholesterol through the cholesteryl ester transfer protein gene does not associate with risk of Alzheimer's disease. Alzheimers Dement (Amst). 2018 Sep 22;10:595-598. eCollection 2018. PubMed PMID: 30422133; PubMed Central PMCID: PMC6215982.
87. Patel D, Mez J, Vardarajan BN, Staley L, Chung J, Zhang X, Farrell JJ, Rynkiewicz MJ, Cannon-Albright LA, Teerlink CC, Stevens J, Corcoran C, Gonzalez Murcia JD, Lopez OL, Mayeux R, Haines JL, Pericak-Vance MA, Schellenberg G, Kauwe JSK, Lunetta KL, Farrer LA; Alzheimer’s Disease Sequencing Project. Association of Rare Coding Mutations With Alzheimer Disease and Other Dementias Among Adults of European Ancestry. JAMA Netw Open. 2019 Mar 1;2(3):e191350. PubMed PMID: 30924900; PubMed Central PMCID: PMC6450321.
88. Raghavan NS, Vardarajan B, Mayeux R. Genomic variation in educational attainment modifies Alzheimer disease risk. Neurol Genet. 2019 Feb 11;5(2):e310. PubMed PMID: 30863791; PubMed Central PMCID: PMC6395060.
89. Raghavan NS, Brickman AM, Andrews H, Manly JJ, Schupf N, Lantigua R, Wolock CJ, Kamalakaran S, Petrovski S, Tosto G, Vardarajan BN, Goldstein DB, Mayeux R; Alzheimer's Disease Sequencing Project. Whole-exome sequencing in 20,197 persons for rare variants in Alzheimer's disease. Ann Clin Transl Neurol. 2018 May 24;5(7):832-842. Erratum in: Ann Clin Transl Neurol. 2019 Feb 25;6(2):416. PubMed PMID: 30009200; PubMed Central PMCID: PMC6043775.
90. Rehker J, Rodhe J, Nesbitt RR, Boyle EA, Martin BK, Lord J, Karaca I, Naj A, Jessen F, Helisalmi S, Soininen H, Hiltunen M, Ramirez A, Scherer M, Farrer LA, Haines JL, Pericak-Vance MA, Raskind WH, Cruchaga C, Schellenberg GD, Joseph B, Brkanac Z. Caspase-8, association with Alzheimer's Disease and functional analysis of rare variants. PLoS One. 2017 Oct 6;12(10):e0185777.. PubMed PMID: 28985224; PubMed Central PMCID: PMC5630132.
91. Reiman EM, Arboleda-Velasquez JF, Quiroz YT, Huentelman MJ, Beach TG, Caselli RJ, Chen Y, Su Y, Myers AJ, Hardy J, Paul Vonsattel J, Younkin SG, Bennett DA, De Jager PL, Larson EB, Crane PK, Keene CD, Kamboh MI, Kofler JK, Duque L, Gilbert JR, Gwirtsman HE, Buxbaum JD, Dickson DW, Frosch MP, Ghetti BF, Lunetta KL, Wang LS, Hyman BT, Kukull WA, Foroud T, Haines JL, Mayeux RP, Pericak-Vance MA, Schneider JA, Trojanowski JQ, Farrer LA, Schellenberg GD, Beecham GW, Montine TJ, Jun GR; Alzheimer’s Disease Genetics Consortium. Exceptionally low likelihood of Alzheimer's dementia in APOE2 homozygotes from a 5,000-person neuropathological study. Nat Commun. 2020 Feb 3;11(1):667. PMID: 32015339; PMCID: PMC6997393.
92. Reitz C, Tosto G, Mayeux R, Luchsinger JA; NIA-LOAD/NCRAD Family Study Group; Alzheimer's Disease Neuroimaging Initiative. Genetic variants in the Fat and Obesity Associated (FTO) gene and risk of Alzheimer's disease. PLoS One. 2012;7(12):e50354. PubMed PMID: 23251365; PubMed Central PMCID: PMC3520931.
93. Reitz C, Lee JH, Rogers RS, Mayeux R. Impact of genetic variation in SORCS1 on memory retention. PLoS One. 2011;6(10):e24588. PubMed PMID: 22046233; PubMed Central PMCID: PMC3202519.
94. Reitz C, Jun G, Naj A, Rajbhandary R, Vardarajan BN, Wang LS, Valladares O, Lin CF, Larson EB, Graff-Radford NR, Evans D, De Jager PL, Crane PK, Buxbaum JD, Murrell JR, Raj T, Ertekin-Taner N, Logue M, Baldwin CT, Green RC, Barnes LL, Cantwell LB, Fallin MD, Go RC, Griffith P, Obisesan TO, Manly JJ, Lunetta KL, Kamboh MI, Lopez OL, Bennett DA, Hendrie H, Hall KS, Goate AM, Byrd GS, Kukull WA, Foroud TM, Haines JL, Farrer LA, Pericak-Vance MA, Schellenberg GD, Mayeux R; Alzheimer Disease Genetics Consortium. Variants in the ATP-binding cassette transporter (ABCA7), apolipoprotein E ϵ4,and the risk of late-onset Alzheimer disease in African Americans. JAMA. 2013 Apr 10;309(14):1483-92. PubMed PMID: 23571587; PubMed Central PMCID: PMC3667653.
95. Reitz C, Mayeux R; Alzheimer’s Disease Genetics Consortium. TREM2 and neurodegenerative disease. N Engl J Med. 2013 Oct 17;369(16):1564-5. PubMed PMID: 24131184; PubMed Central PMCID: PMC3980568.
96. Reitz C, Tosto G, Mayeux R, Luchsinger JA; NIA-LOAD/NCRAD Family Study Group; Alzheimer's Disease Neuroimaging Initiative. Genetic variants in the Fat and Obesity Associated (FTO) gene and risk of Alzheimer's disease. PLoS One. 2012;7(12):e50354. PubMed PMID: 23251365; PubMed Central PMCID: PMC3520931.
97. Reitz C, Tosto G, Mayeux R, Luchsinger JA; NIA-LOAD/NCRAD Family Study Group; Alzheimer's Disease Neuroimaging Initiative. Genetic variants in the Fat and Obesity Associated (FTO) gene and risk of Alzheimer's disease. PLoS One. 2012;7(12):e50354. PubMed PMID: 23251365; PubMed Central PMCID: PMC3520931.
98. Reitz C, Tosto G, Vardarajan B, Rogaeva E, Ghani M, Rogers RS, Conrad C, Haines JL, Pericak-Vance MA, Fallin MD, Foroud T, Farrer LA, Schellenberg GD, George-Hyslop PS, Mayeux R; Alzheimer's Disease Genetics Consortium (ADGC). Independent and epistatic effects of variants in VPS10-d receptors on Alzheimer disease risk and processing of the amyloid precursor protein (APP). Transl Psychiatry. 2013 May 14;3:e256. PubMed PMID: 23673467; PubMed Central PMCID: PMC3669917.
99. Ridge PG, Mukherjee S, Crane PK, Kauwe JS; Alzheimer’s Disease Genetics Consortium. Alzheimer's disease: analyzing the missing heritability. PLoS One. 2013 Nov 7;8(11):e79771. PubMed PMID: 24244562; PubMed Central PMCID: PMC3820606.
100. Ruiz A, Heilmann S, Becker T, Hernández I, Wagner H, Thelen M, Mauleón A, Rosende-Roca M, Bellenguez C, Bis JC, Harold D, Gerrish A, Sims R, Sotolongo-Grau O, Espinosa A, Alegret M, Arrieta JL, Lacour A, Leber M, Becker J, Lafuente A, Ruiz S, Vargas L, Rodríguez O, Ortega G, Dominguez MA; IGAP, Mayeux R, Haines JL, Pericak-Vance MA, Farrer LA, Schellenberg GD, Chouraki V, Launer LJ, van Duijn C, Seshadri S, Antúnez C, Breteler MM, Serrano-Ríos M, Jessen F, Tárraga L, Nöthen MM, Maier W, Boada M, Ramírez A. Follow-up of loci from the International Genomics of Alzheimer's Disease Project identifies TRIP4 as a novel susceptibility gene. Transl Psychiatry. 2014 Feb 4;4:e358. PubMed PMID: 24495969; PubMed Central PMCID: PMC3944635.
101. Shah C, DeMichele-Sweet MA, Sweet RA. Genetics of psychosis of Alzheimer’s disease. Am J Med Genet B Neuropsychiatr Genet. 2017 Jan;174(1):27-35. PMID: 26756273.
102. Sherva R, Gross A, Mukherjee S, Koesterer R, Amouyel P, Bellenguez C, Dufouil C, Bennett DA, Chibnik L, Cruchaga C, Del-Aguila J, Farrer LA, Mayeux R, Munsie L, Winslow A, Newhouse S, Saykin AJ, Kauwe JSK; Alzheimer's Disease Genetics Consortium, Crane PK, Green RC. Genome-wide association study of rate of cognitive decline in Alzheimer’s disease patients identifies novel genes and pathways. Alzheimers Dement. 2020 Aug;16(8):1134-1145. PMID: 32573913
103. Sims R, van der Lee SJ, Naj AC, Bellenguez C, Badarinarayan N, Jakobsdottir J, Kunkle BW, Boland A, Raybould R, Bis JC, Martin ER, Grenier-Boley B, Heilmann-Heimbach S, Chouraki V, Kuzma AB, Sleegers K, Vronskaya M, Ruiz A, Graham RR, Olaso R, Hoffmann P, Grove ML, Vardarajan BN, Hiltunen M, Nöthen MM, White CC, Hamilton-Nelson KL, Epelbaum J, Maier W, Choi SH, Beecham GW, Dulary C, Herms S, Smith AV, Funk CC, Derbois C, Forstner AJ, Ahmad S, Li H, Bacq D, Harold D, Satizabal CL, Valladares O, Squassina A, Thomas R, Brody JA, Qu L, Sánchez-Juan P, Morgan T, Wolters FJ, Zhao Y, Garcia FS, Denning N, Fornage M, Malamon J, Naranjo MCD, Majounie E, Mosley TH, Dombroski B, Wallon D, Lupton MK, Dupuis J, Whitehead P, Fratiglioni L, Medway C, Jian X, Mukherjee S, Keller L, Brown K, Lin H, Cantwell LB, Panza F, McGuinness B, Moreno-Grau S, Burgess JD, Solfrizzi V, Proitsi P, Adams HH, Allen M, Seripa D, Pastor P, Cupples LA, Price ND, Hannequin D, Frank-García A, Levy D, Chakrabarty P, Caffarra P, Giegling I, Beiser AS, Giedraitis V, Hampel H, Garcia ME, Wang X, Lannfelt L, Mecocci P, Eiriksdottir G, Crane PK, Pasquier F, Boccardi V, Henández I, Barber RC, Scherer M, Tarraga L, Adams PM, Leber M, Chen Y, Albert MS, Riedel-Heller S, Emilsson V, Beekly D, Braae A, Schmidt R, Blacker D, Masullo C, Schmidt H, Doody RS, Spalletta G, Longstreth WT Jr, Fairchild TJ, Bossù P, Lopez OL, Frosch MP, Sacchinelli E, Ghetti B, Yang Q, Huebinger RM, Jessen F, Li S, Kamboh MI, Morris J, Sotolongo-Grau O, Katz MJ, Corcoran C, Dunstan M, Braddel A, Thomas C, Meggy A, Marshall R, Gerrish A, Chapman J, Aguilar M, Taylor S, Hill M, Fairén MD, Hodges A, Vellas B, Soininen H, Kloszewska I, Daniilidou M, Uphill J, Patel Y, Hughes JT, Lord J, Turton J, Hartmann AM, Cecchetti R, Fenoglio C, Serpente M, Arcaro M, Caltagirone C, Orfei MD, Ciaramella A, Pichler S, Mayhaus M, Gu W, Lleó A, Fortea J, Blesa R, Barber IS, Brookes K, Cupidi C, Maletta RG, Carrell D, Sorbi S, Moebus S, Urbano M, Pilotto A, Kornhuber J, Bosco P, Todd S, Craig D, Johnston J, Gill M, Lawlor B, Lynch A, Fox NC, Hardy J; ARUK Consortium, Albin RL, Apostolova LG, Arnold SE, Asthana S, Atwood CS, Baldwin CT, Barnes LL, Barral S, Beach TG, Becker JT, Bigio EH, Bird TD, Boeve BF, Bowen JD, Boxer A, Burke JR, Burns JM, Buxbaum JD, Cairns NJ, Cao C, Carlson CS, Carlsson CM, Carney RM, Carrasquillo MM, Carroll SL, Diaz CC, Chui HC, Clark DG, Cribbs DH, Crocco EA, DeCarli C, Dick M, Duara R, Evans DA, Faber KM, Fallon KB, Fardo DW, Farlow MR, Ferris S, Foroud TM, Galasko DR, Gearing M, Geschwind DH, Gilbert JR, Graff-Radford NR, Green RC, Growdon JH, Hamilton RL, Harrell LE, Honig LS, Huentelman MJ, Hulette CM, Hyman BT, Jarvik GP, Abner E, Jin LW, Jun G, Karydas A, Kaye JA, Kim R, Kowall NW, Kramer JH, LaFerla FM, Lah JJ, Leverenz JB, Levey AI, Li G, Lieberman AP, Lunetta KL, Lyketsos CG, Marson DC, Martiniuk F, Mash DC, Masliah E, McCormick WC, McCurry SM, McDavid AN, McKee AC, Mesulam M, Miller BL, Miller CA, Miller JW, Morris JC, Murrell JR, Myers AJ, O'Bryant S, Olichney JM, Pankratz VS, Parisi JE, Paulson HL, Perry W, Peskind E, Pierce A, Poon WW, Potter H, Quinn JF, Raj A, Raskind M, Reisberg B, Reitz C, Ringman JM, Roberson ED, Rogaeva E, Rosen HJ, Rosenberg RN, Sager MA, Saykin AJ, Schneider JA, Schneider LS, Seeley WW, Smith AG, Sonnen JA, Spina S, Stern RA, Swerdlow RH, Tanzi RE, Thornton-Wells TA, Trojanowski JQ, Troncoso JC, Van Deerlin VM, Van Eldik LJ, Vinters HV, Vonsattel JP, Weintraub S, Welsh-Bohmer KA, Wilhelmsen KC, Williamson J, Wingo TS, Woltjer RL, Wright CB, Yu CE, Yu L, Garzia F, Golamaully F, Septier G, Engelborghs S, Vandenberghe R, De Deyn PP, Fernadez CM, Benito YA, Thonberg H, Forsell C, Lilius L, Kinhult-Stählbom A, Kilander L, Brundin R, Concari L, Helisalmi S, Koivisto AM, Haapasalo A, Dermecourt V, Fievet N, Hanon O, Dufouil C, Brice A, Ritchie K, Dubois B, Himali JJ, Keene CD, Tschanz J, Fitzpatrick AL, Kukull WA, Norton M, Aspelund T, Larson EB, Munger R, Rotter JI, Lipton RB, Bullido MJ, Hofman A, Montine TJ, Coto E, Boerwinkle E, Petersen RC, Alvarez V, Rivadeneira F, Reiman EM, Gallo M, O'Donnell CJ, Reisch JS, Bruni AC, Royall DR, Dichgans M, Sano M, Galimberti D, St George-Hyslop P, Scarpini E, Tsuang DW, Mancuso M, Bonuccelli U, Winslow AR, Daniele A, Wu CK; GERAD/PERADES, CHARGE, ADGC, EADI, Peters O, Nacmias B, Riemenschneider M, Heun R, Brayne C, Rubinsztein DC, Bras J, Guerreiro R, Al-Chalabi A, Shaw CE, Collinge J, Mann D, Tsolaki M, Clarimón J, Sussams R, Lovestone S, O'Donovan MC, Owen MJ, Behrens TW, Mead S, Goate AM, Uitterlinden AG, Holmes C, Cruchaga C, Ingelsson M, Bennett DA, Powell J, Golde TE, Graff C, De Jager PL, Morgan K, Ertekin-Taner N, Combarros O, Psaty BM, Passmore P, Younkin SG, Berr C, Gudnason V, Rujescu D, Dickson DW, Dartigues JF, DeStefano AL, Ortega-Cubero S, Hakonarson H, Campion D, Boada M, Kauwe JK, Farrer LA, Van Broeckhoven C, Ikram MA, Jones L, Haines JL, Tzourio C, Launer LJ, Escott-Price V, Mayeux R, Deleuze JF, Amin N, Holmans PA, Pericak-Vance MA, Amouyel P, van Duijn CM, Ramirez A, Wang LS, Lambert JC, Seshadri S, Williams J, Schellenberg GD. Rare coding variants in PLCG2, ABI3, and TREM2 implicate microglial-mediated innate immunity in Alzheimer's disease. Nat Genet. 2017 Sep;49(9):1373-1384. PubMed PMID: 28714976; PubMed Central PMCID: PMC5669039.
104. Swaminathan S, Huentelman MJ, Corneveaux JJ, Myers AJ, Faber KM, Foroud T, Mayeux R, Shen L, Kim S, Turk M, Hardy J, Reiman EM, Saykin AJ; Alzheimer's Disease Neuroimaging Initiative and NIA-LOAD/NCRAD Family Study Group. Analysis of copy number variation in Alzheimer's disease in a cohort of clinically characterized and neuropathologically verified individuals. PLoS One. 2012;7(12):e50640. PubMed PMID: 23227193; PubMed Central PMCID: PMC3515604.
105. Swaminathan S, Shen L, Kim S, Inlow M, West JD, Faber KM, Foroud T, Mayeux R, Saykin AJ; Alzheimer's Disease Neuroimaging Initiative; NIA-LOAD/NCRAD Family Study Group. Analysis of copy number variation in Alzheimer's disease: the NIALOAD/ NCRAD Family Study. Curr Alzheimer Res. 2012 Sep;9(7):801-14. PubMed PMID: 22486522; PubMed Central PMCID: PMC3500615.
106. Sweet RA, Bennett DA, Graff-Radford NR, Mayeux R; National Institute on Aging Late-Onset Alzheimer's Disease Family Study Group. Assessment and familial aggregation of psychosis in Alzheimer's disease from the National Institute on Aging Late Onset Alzheimer's Disease Family Study. Brain. 2010 Apr;133(Pt 4):1155-62. PubMed PMID: 20147454; PubMed Central PMCID: PMC2912688.
107. Tan CH, Fan CC, Mormino EC, Sugrue LP, Broce IJ, Hess CP, Dillon WP, Bonham LW, Yokoyama JS, Karch CM, Brewer JB, Rabinovici GD, Miller BL, Schellenberg GD, Kauppi K, Feldman HA, Holland D, McEvoy LK, Hyman BT, Bennett DA, Andreassen OA, Dale AM, Desikan RS; Alzheimer’s Disease Neuroimaging Initiative. Polygenic hazard score: an enrichment marker for Alzheimer's associated amyloid and tau deposition. Acta Neuropathol. 2018 Jan;135(1):85-93. PubMed PMID: 29177679; PubMed Central PMCID: PMC5758038.
108. Tan CH, Hyman BT, Tan JJX, Hess CP, Dillon WP, Schellenberg GD, Besser LM, Kukull WA, Kauppi K, McEvoy LK, Andreassen OA, Dale AM, Fan CC, Desikan RS. Polygenic hazard scores in preclinical Alzheimer disease. Ann Neurol. 2017 Sep;82(3):484-488. PubMed PMID: 28940650; PubMed Central PMCID: PMC5758043.
109. Teslovich TM, Kim DS, Yin X, Stancáková A, Jackson AU, Wielscher M, Naj A, Perry JRB, Huyghe JR, Stringham HM, Davis JP, Raulerson CK, Welch RP, Fuchsberger C, Locke AE, Sim X, Chines PS, Narisu N, Kangas AJ, Soininen P; Genetics of Obesity-Related Liver Disease Consortium (GOLD), The Alzheimer's Disease Genetics Consortium (ADGC), The DIAbetes Genetics Replication And Meta-analysis (DIAGRAM), Ala-Korpela M, Gudnason V, Musani SK, Jarvelin MR, Schellenberg GD, Speliotes EK, Kuusisto J, Collins FS, Boehnke M, Laakso M, Mohlke KL.Identification of seven novel loci associated with amino acid levels using single-variant and gene-based tests in 8545 Finnish men from the METSIM study. Hum Mol Genet. 2018 May 1;27(9):1664-1674. PMID: 29481666
110. Tosto G, Bird TD, Bennett DA, Boeve BF, Brickman AM, Cruchaga C, Faber K, Foroud TM, Farlow M, Goate AM, Graff-Radford NR, Lantigua R, Manly J, Ottman R, Rosenberg R, Schaid DJ, Schupf N, Stern Y, Sweet RA, Mayeux R; National Institute on Aging Late-Onset Alzheimer Disease/National Cell Repository for Alzheimer Disease (NIA-LOAD/NCRAD) Family Study Group. The Role of Cardiovascular Risk Factors and Stroke in Familial Alzheimer Disease. JAMA Neurol. 2016 Oct 1;73(10):1231-1237. PubMed PMID: 27533593; PubMed Central PMCID: PMC5155512.
111. Tosto G, Bird TD, Tsuang D, Bennett DA, Boeve BF, Cruchaga C, Faber K, Foroud TM, Farlow M, Goate AM, Bertlesen S, Graff-Radford NR, Medrano M, Lantigua R, Manly J, Ottman R, Rosenberg R, Schaid DJ, Schupf N, Stern Y, Sweet RA, Mayeux R. Polygenic risk scores in familial Alzheimer disease. Neurology. 2017 Mar 21;88(12):1180-1186. PubMed PMID: 28213371; PubMed Central PMCID: PMC5373783.
112. Tosto G, Fu H, Vardarajan BN, Lee JH, Cheng R, Reyes-Dumeyer D, Lantigua R, Medrano M, Jimenez-Velazquez IZ, Elkind MS, Wright CB, Sacco RL, Pericak-Vance M, Farrer L, Rogaeva E, St George-Hyslop P, Reitz C, Mayeux R. F-box/LRR-repeat protein 7 is genetically associated with Alzheimer's disease. Ann Clin Transl Neurol. 2015 Aug;2(8):810-20. PubMed PMID: 26339675; PubMed Central PMCID: PMC4554442.
113. Tosto G, Bird TD, Tsuang D, Bennett DA, Boeve BF, Cruchaga C, Faber K, Foroud TM, Farlow M, Goate AM, Bertlesen S, Graff-Radford NR, Medrano M, Lantigua R, Manly J, Ottman R, Rosenberg R, Schaid DJ, Schupf N, Stern Y, Sweet RA, Mayeux R. Polygenic risk scores in familial Alzheimer disease. Neurology. 2017 Mar 21;88(12):1180-1186. PubMed PMID: 28213371; PubMed Central PMCID: PMC5373783.
114. Tosto G, Vardarajan B, Sariya S, Brickman AM, Andrews H, Manly JJ, Schupf N, Reyes-Dumeyer D, Lantigua R, Bennett DA, De Jager PL, Mayeux R. Association of Variants in PINX1 and TREM2 With Late-Onset Alzheimer Disease. JAMA Neurol. 2019 May 6. PubMed PMID: 31058951; PubMed Central PMCID: PMC6503572.
115. Tsuang D, Leverenz JB, Lopez OL, Hamilton RL, Bennett DA, Schneider JA, Buchman AS, Larson EB, Crane PK, Kaye JA, Kramer P, Woltjer R, Trojanowski JQ, Weintraub D, Chen-Plotkin AS, Irwin DJ, Rick J, Schellenberg GD, Watson GS, Kukull W, Nelson PT, Jicha GA, Neltner JH, Galasko D, Masliah E, Quinn JF, Chung KA, Yearout D, Mata IF, Wan JY, Edwards KL, Montine TJ, Zabetian CP. APOE ε4 increases risk for dementia in pure synucleinopathies. JAMA Neurol. 2013 Feb;70(2):223-8. PubMed PMID: 23407718; PubMed Central PMCID: PMC3580799.
116. Vardarajan BN, Faber KM, Bird TD, Bennett DA, Rosenberg R, Boeve BF, Graff-Radford NR, Goate AM, Farlow M, Sweet RA, Lantigua R, Medrano MZ, Ottman R, Schaid DJ, Foroud TM, Mayeux R; NIA-LOAD/NCRAD Family Study Group. Age-specific incidence rates for dementia and Alzheimer disease in NIA-LOAD/NCRAD and EFIGA families: National Institute on Aging Genetics Initiative for Late-Onset Alzheimer Disease/National Cell Repository for Alzheimer Disease (NIA-LOAD/NCRAD) and Estudio Familiar de Influencia Genetica en Alzheimer (EFIGA). JAMA Neurol. 2014 Mar;71(3):315-23. PubMed PMID: 24425039; PubMed Central PMCID: PMC4000602.
117. Vardarajan BN, Tosto G, Lefort R, Yu L, Bennett DA, De Jager PL, Barral S, Reyes-Dumeyer D, Nagy PL, Lee JH, Cheng R, Medrano M, Lantigua R, Rogaeva E, St George-Hyslop P, Mayeux R. Ultra-rare mutations in SRCAP segregate in Caribbean Hispanic families with Alzheimer disease. Neurol Genet. 2017 Aug 24;3(5):e178. PubMed PMID: 28852706; PubMed Central PMCID: PMC5570674.
118. Wang LS, Naj AC, Graham RR, Crane PK, Kunkle BW, Cruchaga C, Murcia JD, Cannon-Albright L, Baldwin CT, Zetterberg H, Blennow K, Kukull WA, Faber KM, Schupf N, Norton MC, Tschanz JT, Munger RG, Corcoran CD, Rogaeva E; Alzheimer's Disease Genetics Consortium, Lin CF, Dombroski BA, Cantwell LB, Partch A, Valladares O, Hakonarson H, St George-Hyslop P, Green RC, Goate AM, Foroud TM, Carney RM, Larson EB, Behrens TW, Kauwe JS, Haines JL, Farrer LA, Pericak-Vance MA, Mayeux R, Schellenberg GD; National Institute on Aging-Late-Onset Alzheimer’s Disease (NIA-LOAD) Family Study, Albert MS, Albin RL, Apostolova LG, Arnold SE, Barber R, Barmada M, Barnes LL, Beach TG, Becker JT, Beecham GW, Beekly D, Bennett DA, Bigio EH, Bird TD, Blacker D, Boeve BF, Bowen JD, Boxer A, Burke JR, Buxbaum JD, Cairns NJ, Cao C, Carlson CS, Carroll SL, Chui HC, Clark DG, Cribbs DH, Crocco EA, DeCarli C, DeKosky ST, Demirci FY, Dick M, Dickson DW, Duara R, Ertekin-Taner N, Fallon KB, Farlow MR, Ferris S, Frosch MP, Galasko DR, Ganguli M, Gearing M, Geschwind DH, Ghetti B, Gilbert JR, Glass JD, Graff-Radford NR, Growdon JH, Hamilton RL, Hamilton-Nelson KL, Harrell LE, Head E, Honig LS, Hulette CM, Hyman BT, Jarvik GP, Jicha GA, Jin LW, Jun G, Jun G, Kamboh MI, Karydas A, Kaye JA, Kim R, Koo EH, Kowall NW, Kramer JH, LaFerla FM, Lah JJ, Leverenz JB, Levey AI, Li G, Lieberman AP, Lopez OL, Lunetta KL, Lyketsos CG, Mack WJ, Marson DC, Martin ER, Martiniuk F, Mash DC, Masliah E, McCormick WC, McCurry SM, McDavid AN, McKee AC, Mesulam WM, Miller BL, Miller CA, Miller JW, Montine TJ, Morris JC, Murrell JR, Olichney JM, Parisi JE, Perry W, Peskind E, Petersen RC, Pierce A, Poon WW, Potter H, Quinn JF, Raj A, Raskind M, Reiman EM, Reisberg B, Reitz C, Ringman JM, Roberson ED, Rosen HJ, Rosenberg RN, Sano M, Saykin AJ, Schneider JA, Schneider LS, Seeley WW, Smith AG, Sonnen JA, Spina S, Stern RA, Tanzi RE, Thornton-Wells TA, Trojanowski JQ, Troncoso JC, Tsuang DW, Van Deerlin VM, Van Eldik LJ, Vardarajan BN, Vinters HV, Vonsattel JP, Weintraub S, Welsh-Bohmer KA, Williamson J, Wishnek S, Woltjer RL, Wright CB, Younkin SG, Yu CE, Yu L. Rarity of the Alzheimer disease-protective APP A673T variant in the United States. JAMA Neurol. 2015 Feb;72(2):209-16. PubMed PMID: 25531812; PubMed Central PMCID: PMC4324097.
119. Wetzel-Smith MK, Hunkapiller J, Bhangale TR, Srinivasan K, Maloney JA, Atwal JK, Sa SM, Yaylaoglu MB, Foreman O, Ortmann W, Rathore N, Hansen DV, Tessier-Lavigne M; Alzheimer's Disease Genetics Consortium, Mayeux R, Pericak-Vance M, Haines J, Farrer LA, Schellenberg GD, Goate A, Behrens TW, Cruchaga C, Watts RJ, Graham RR. A rare mutation in UNC5C predisposes to late-onset Alzheimer's disease and increases neuronal cell death. Nat Med. 2014 Dec;20(12):1452-7. PubMed PMID: 25419706; PubMed Central PMCID: PMC4301587.
120. Wijsman EM, Pankratz ND, Choi Y, Rothstein JH, Faber KM, Cheng R, Lee JH, Bird TD, Bennett DA, Diaz-Arrastia R, Goate AM, Farlow M, Ghetti B, Sweet RA, Foroud TM, Mayeux R; NIA-LOAD/NCRAD Family Study Group. Genome-wide association of familial late-onset Alzheimer's disease replicates BIN1 and CLU and nominates CUGBP2 in interaction with APOE. PLoS Genet. 2011 Feb;7(2):e1001308. PubMed PMID: 21379329; PubMed Central PMCID: PMC3040659.
121. Wijsman EM, Pankratz ND, Choi Y, Rothstein JH, Faber KM, Cheng R, Lee JH, Bird TD, Bennett DA, Diaz-Arrastia R, Goate AM, Farlow M, Ghetti B, Sweet RA, Foroud TM, Mayeux R; NIA-LOAD/NCRAD Family Study Group. Genome-wide association of familial late-onset Alzheimer's disease replicates BIN1 and CLU and nominates CUGBP2 in interaction with APOE. PLoS Genet. 2011 Feb;7(2):e1001308. PubMed PMID: 21379329; PubMed Central PMCID: PMC3040659.
122. Zhang X, Zhu C, Beecham G, Vardarajan BN, Ma Y, Lancour D, Farrell JJ, Chung J; Alzheimer's Disease Sequencing Project, Mayeux R, Haines JL, Schellenberg GD, Pericak-Vance MA, Lunetta KL, Farrer LA. A rare missense variant of CASP7 is associated with familial late-onset Alzheimer's disease. Alzheimers Dement. 2019 Mar;15(3):441-452. PubMed PMID: 30503768; PubMed Central PMCID: PMC6408965.
123. Zhao L, He Z, Zhang D, Wang GT, Renton AE, Vardarajan BN, Nothnagel M, Goate AM, Mayeux R, Leal SM. A Rare Variant Nonparametric Linkage Method for Nuclear and Extended Pedigrees with Application to Late-Onset Alzheimer Disease via WGS Data. Am J Hum Genet. 2019 Oct 3;105(4):822-835. PubMed PMID: 31585107.
124. Zou F, Chai HS, Younkin CS, Allen M, Crook J, Pankratz VS, Carrasquillo MM, Rowley CN, Nair AA, Middha S, Maharjan S, Nguyen T, Ma L, Malphrus KG, Palusak R, Lincoln S, Bisceglio G, Georgescu C, Kouri N, Kolbert CP, Jen J, Haines JL, Mayeux R, Pericak-Vance MA, Farrer LA, Schellenberg GD; Alzheimer's Disease Genetics Consortium, Petersen RC, Graff-Radford NR, Dickson DW, Younkin SG, Ertekin-Taner N. Brain expression genome-wide association study (eGWAS) identifies human disease-associated variants. PLoS Genet. 2012;8(6):e1002707. PubMed PMID: 22685416; PubMed Central PMCID: PMC3369937.
125. Prokopenko D, Morgan SL, Mullin K, Hofmann O, Chapman B, Kirchner R, Alzheimer’s Disease Neuroimaging Initiative, Amberkar S, Wohlers I, Lange C, Hide W, Bertram L, Tanzi RE. Whole‐genome sequencing reveals new Alzheimer's disease–associated rare variants in loci related to synaptic function and neuronal development. Alzheimer’s Dement. 2021; 1-19.
